# Patterns of depression prevalence and antidepressant use in South Africa, 2002-2024: a system dynamics modelling perspective

**DOI:** 10.64898/2026.03.08.26347878

**Authors:** Leigh F. Johnson, Danielle Giovenco, Katherine Eyal, Ashleigh Craig, Inge Petersen, Mpho Tlali, Naomi Levitt, Max Bachmann, Andreas D. Haas, Lara Fairall

## Abstract

**Background:** Depression is estimated to be the leading cause of disability in South Africa, yet data on depression prevalence and antidepressant use are inconsistent and fragmentary. We present a system dynamics modelling approach to integrate these data and assess trends and inequalities in depression prevalence and treatment access.

**Methods:** We developed a deterministic model of the South African population aged 15 and older, stratified by age, sex, HIV status/stage and susceptibility to depression. Individual transitions between depressed/healthy and treated/untreated states were simulated over time, from 1985. The model was calibrated to depression prevalence data from nine nationally representative household surveys (2002-2024) and ten smaller studies reporting prevalence of antidepressant use, using a Bayesian approach.

**Results:** The model estimated a slight decline in depression point prevalence over time, from 5.1% (95% CI: 4.5-5.6%) in 2002 to 4.5% (95% CI: 4.0-5.0%) in 2024, although with a transient rise in depression prevalence during the COVID-19 period. In 2024, depression prevalence was higher in women (5.3%, 95% CI: 4.7-5.9%) than men (3.6%, 95% CI: 3.2-4.0%), and highest at ages 60 and older. The lifetime prevalence of depression was 70.6% (95% CI: 67.8-73.6); alternative model settings with a more concentrated distribution of depression risk were inconsistent with longitudinal data. The proportion of adults using antidepressants increased from 1.0% (95% CI: 0.8-1.2%) in 2008 to 2.8% (95% CI: 2.2-3.4%) in 2024. In 2024, antidepressant use was 11.0% (95% CI: 8.8-13.5%) in the private sector, compared to only 0.9% (95% CI: 0.7-1.1%) in the rest of the population, and the ratio of new antidepressant initiations to new cases of depression was 0.12 nationally.

**Conclusion:** The prevalence of depression in South Africa has been relatively stable over the last two decades. Although antidepressant use has increased, overall use remains low, and substantial inequality remains in access to treatment in the public and private health sectors.

## Introduction

Depression is estimated to be the second leading cause of years lived with disability globally [1], with sub-Saharan Africa being the region with the highest prevalence [2]. Antidepressants have the potential to alleviate depression symptoms [3] and prevent recurrences [4, 5]. In Western Europe, Canada, the US and Australia, the proportion of the population using antidepressants varies between 4 and 16% [6–9], with rising use over the last two decades [8–11]. However, little is known about the prevalence of antidepressant use in low- and middle-income countries like South Africa [6].

Depression is usually episodic, with symptoms typically lasting for several months [12, 13]. Cross-sectional surveys have estimated that the lifetime prevalence of depression globally is around 10-15% [14, 15], based on retrospective self-report. This has supported the view that only a small fraction of the population is at risk of depression, and that this sub-population experiences high rates of recurrence, with the disease assuming characteristics of a ‘chronic condition’ [16]. However, a number of studies have noted methodological problems with these retrospective lifetime prevalence estimates, most importantly failure to recall past depression episodes [17–21]. These studies estimate that the cumulative lifetime risk is much greater than 15%, probably closer to 50% [22]. This would imply that a much larger fraction of the population is at risk of depression, although many experience only a single episode, with low recurrence risk. Distinguishing depression patients at high and low risk of recurrence is potentially important for ensuring cost-effective allocation of mental healthcare resources, but there have been few successful attempts to address this challenge [23].

South Africa is a middle-income country, in which mental disorders are estimated to be the leading contributor to years lived with disability [24], as well as causing over $3 billion per annum in lost productivity [25]. Several known risk factors for depression are highly prevalent, including exposure to violence, heavy alcohol use, human immunodeficiency virus (HIV), other chronic conditions, poverty and adverse childhood experiences [26, 27]. The National Mental Health Policy Framework and Strategic Plan recognizes the importance of addressing these risk factors [28], but the overall prevalence of depression is poorly quantified. A number of nationally representative surveys have estimated the prevalence of depression [29–31], but only one, conducted between 2002 and 2004, used a diagnostic interview [29]; the other surveys have all used screening instruments for depressive symptoms [30, 31], which are likely to overstate the true prevalence of depression [32]. The National Mental Health Policy Framework proposes several indicators to track the implementation of policy [28], but there are almost no indicators for tracking the overall mental health disease burden, and hence the impact of policy changes [33].

Mathematical models are important tools to support health policy: they can be used to estimate the magnitude of a health problem, to evaluate the performance of current health programmes, to predict the potential impact of changes in these programmes, and to test hypotheses about disease processes [34]. Models of depression vary in complexity, from simple tools to estimate the prevalence or incidence of depression (such as used in the Global Burden of Disease study [35]) to more complex system dynamics models that simulate interactions between individual-and population-level processes [36] and the social determinants of depression [37]. In the context of these system dynamics models, Bayesian evidence synthesis is a powerful tool for integrating different types of data to understand disease trends and to assess the plausibility of different models [38]. Models have previously been used to demonstrate the extreme heterogeneity in depression recurrence risk [39] and the likely biases in lifetime prevalence estimates [18, 21]. However, there have been relatively few attempts to develop simulation models in mental health to support population-level decision making [36], especially in resource-limited settings, where infectious diseases are often viewed as more pressing health problems.

This study aims to synthesize different South African datasets, in order to estimate trends and inequalities in the prevalence of depression and antidepressant use. We do this by developing a system dynamics model of depression and calibrating it to published data, using a Bayesian framework. We further aim to use this model to evaluate the plausibility of different assumptions about heterogeneity in depression incidence.

## Methods

### Model structure

The model is compartmental and deterministic. The adult South African population is divided into compartments defined by age (in single years of age from 15 to 90+), sex and HIV status/stage (HIV-negative, HIV-positive undiagnosed, HIV-diagnosed but untreated, and on antiretroviral treatment [ART]). The numbers in each age/sex/HIV compartment, in each year, are fixed inputs from the Thembisa model, an integrated HIV and demographic model developed for South Africa [40]. The population is further divided into individuals at high and low risk of depression, with the high-risk group defined as individuals who are at high risk of depression and recurrence (due to genetics, biochemical factors and/or adverse childhood experiences) and the low-risk group representing individuals who may experience depression but who are unlikely to experience recurrences. Within each of these compartments, individuals are divided into a number of additional compartments defined by their depression disease state and treatment history (Figure 1).

**Figure 1:**
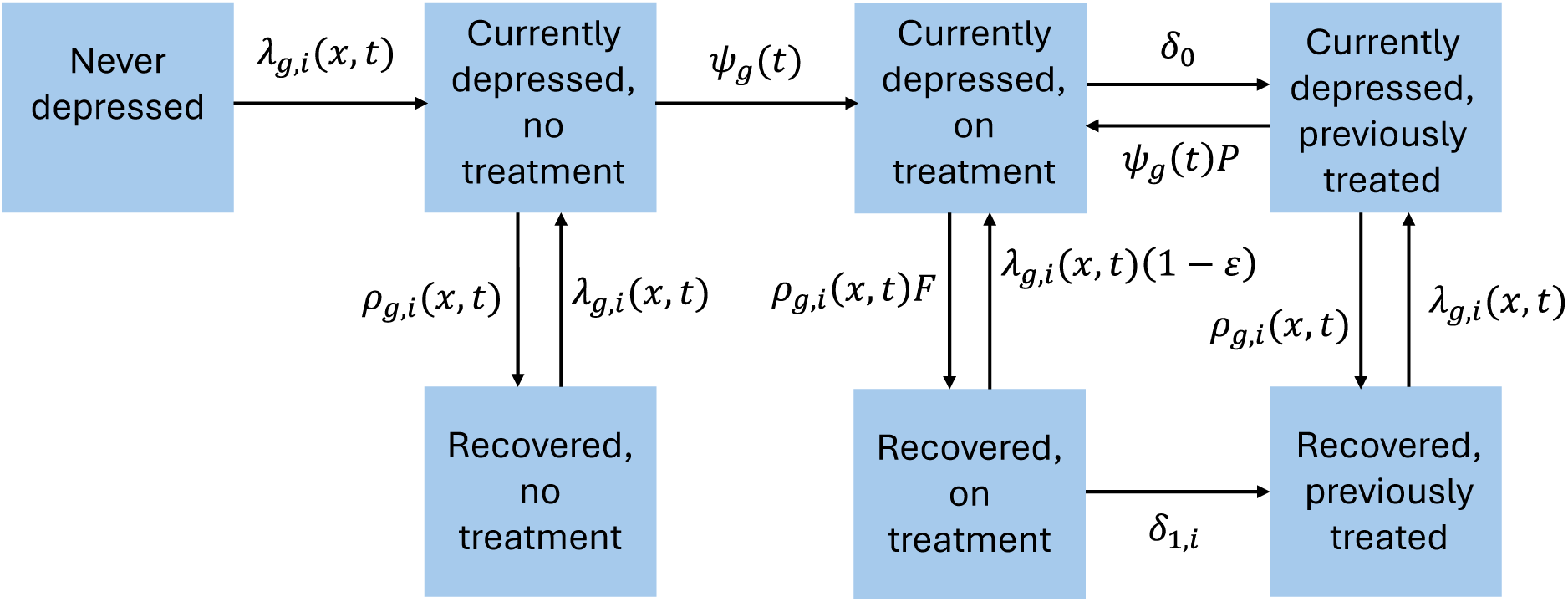
Model of depression incidence, treatment and resolution. The states shown are replicated for all possible combinations of age (𝑥), sex (𝑔), depression risk group (𝑖) and HIV status/stage.

The numbers of individuals in each model compartment are updated at weekly time steps, starting from mid-1985. The model makes allowance for a number of risk factors identified in the National Mental Health Policy Framework [28] and a recent umbrella review [26], which are likely to be important in explaining age, sex and temporal variation in depression incidence.

For an HIV-negative individual of sex 𝑔 and age 𝑥, in risk group 𝑖, the annual rate of depression incidence in year 𝑡, 𝜆_𝑔,𝑖_(𝑥, 𝑡), is calculated as

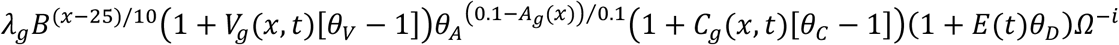

where 𝜆_𝑔_ is the depression incidence in low-risk individuals with no risk factors at age 25; 𝐵 is the factor by which depression risk increases per 10 years of age; 𝑉_𝑔_(𝑥, 𝑡) is the proportion of individuals who have experienced physical violence in the last 12 months; 𝜃_𝑉_ is the relative risk of depression in people who have experienced physical violence in the last year; 𝜃_𝐴_ is the relative risk of depression per 10% reduction in binge drinking prevalence; 𝐴_𝑔_(𝑥) is the prevalence of binge drinking; 𝐶_𝑔_(𝑥, 𝑡) is the prevalence of chronic conditions that are associated with increased depression risk (arthritis, asthma, ischaemic heart disease, stroke); 𝜃_𝐶_ is the relative risk of depression in people with these chronic conditions; 𝐸(𝑡) is the excess monthly mortality per 100 000 population during the COVID-19 pandemic period; 𝜃_𝐷_is the proportional increase in depression incidence per unit increase in excess deaths (per month, per 100 000 population); and 𝛺 is the relative incidence of depression incidence in low-risk individuals (𝑖=0) relative to high-risk individuals (𝑖=1). For HIV-positive individuals, the depression incidence is further increased by a factor 𝐻_1_ if they are diagnosed but untreated or 𝐻_2_ if they are treated or undiagnosed. The values of these parameters are summarized in Table 1 and more detailed explanations are provided in section 1.1 of the supplementary materials.

**Table 1:** Key model parameters.

| Parameter | Symbol | Mean | SD* | Sources |
| --- | --- | --- | --- | --- |
| **Structural parameters (Figure 1)** |  |  |  |  |
| Proportion of adults in high-risk group | - | 0.07 | - | [39, 41] |
| Incidence of depression in low-risk men | $\lambda_0$ | 0.027 | 0.010 | [32] |
| Incidence of depression in low-risk women | $\lambda_1$ | 0.034 | 0.013 | [32] |
| Ratio of incidence in low-risk group to high-risk group | $\Omega$ | 0.30 | 0.20 | - |
| Increase in depression incidence per 10-year increase in age (RR) | $B$ | 1 | 0.10 | - |
| RR of depression in people who experienced violence | $\theta_V$ | 1.56 | - | QoL survey 2020-21 |
| RR of depression per 10% reduction in the prevalence of binge drinking | $\theta_A$ | 0.96 | - | [42, 43] |
| RR of depression in diagnosed people living with HIV |  |  |  |  |
| Untreated | $H_1$ | 2.00 | - | [27, 44-46] |
| Treated | $H_2$ | 1.20 | - | [47] |
| RR of depression in people with asthma, arthritis, IHD or stroke | $\theta_C$ | 2.5 | - | [47] |
| Increase in depression incidence per unit excess deaths (per 100,000) | $\theta_D$ | 0.039 | 0.01 | [48] |
| Mean duration of symptoms if untreated (months) | $D$ | 9 | 3 | [12, 49] |
| Proportional reduction in symptoms per log increase in MET-minutes/week | $\alpha$ | 0.135 | - | [50] |
| Annual health seeking rate in women | $\gamma$ | 0.2 | 0.1 | [51] |
| RR of health seeking in men | $K$ | 0.3 | 0.15 | [51-53] |
| Proportion prescribed antidepressants pre-2008 | $\eta$ | 0.15 | 0.09 | [53] |
| RR of antidepressants in 2018 | $J(2018)$ | 2 | 0.6 | [53, 54] |
| RR of health seeking if previously treated | $P$ | 9 | 5 | [53] |
| RR of depression recovery if treated | $F$ | 1.8 | - | [3] |
| Reduction in recurrence if treated | $\varepsilon$ | 0.3 | - | [4, 5] |
| Average months on antidepressants while experiencing symptoms | $12/\delta_0$ | 7 | 3 | [55-59] |
| Average years on antidepressants post-recovery | $1/\delta_1$ | 6 | 2 | [52, 59] |
| **Calibration parameters** |  |  |  |  |
| Ratio of point prevalence to 12-month prevalence, using CIDI | $R_1$ | 0.77 | 0.08 | [32] |
| Ratio of true prevalence to PHQ-9 prevalence<sup>†</sup> | $R_2$ | 0.35 | 0.055 | [32] |
| Ratio of true prevalence to CES-D prevalence<sup>†</sup> | $R_3$ | 0.35 | 0.055 | [32] |
| Effect of log(prevalence) on association between initial and later depression | $\beta$ | -0.16 | 0.04 | NIDS data |
| RR of antidepressant use in public sector | $U$ | 0.09 | 0.03 | [53] |
| RR of antidepressant use in people receiving treatment for other chronic conditions | $\chi$ | 3.00 | 1.00 | [60-62] |
| Coefficient of variation in random effects | $\mu$ | 1.00 | 0.29 | - |

CES-D = Centre for Epidemiological Studies Depression Scale. CIDI = Composite International Diagnostic Interview, IHD = ischaemic heart disease, MET = metabolic equivalent of time, NIDS = National Income Dynamics Study [31], PHQ-9 = Patient Health Questionnaire-9, QoL Survey = Quality of Life Survey [63, 64], RR = relative rate, SD = standard deviation. \* Where no standard deviation is specified, the parameter was fixed at the mean value for calibration purposes. † A threshold of 10 or more is used when calculating the prevalence of ‘probable depression’ using the PHQ-9 and CES-D-10 symptom scores.

In the absence of treatment, individuals with depression are assumed to recover at annual rate 𝜌_𝑔,𝑖_(𝑥, 𝑡), which takes into account the level of physical activity [50]. This is calculated as

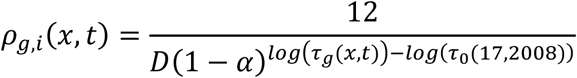

where 𝐷 is the average duration of depression (in months) in a highly active reference category (which we arbitrarily set to 17-year old males in 2008); and 𝛼 is the proportionate reduction in the duration of depression per log increase in 𝜏_𝑔_(𝑥, 𝑡), the metabolic equivalent of time (MET) minutes per week. Parameter values are summarized in Table 1 and explained more fully in section 1.2 of the supplementary materials.

Individuals experiencing depression, who have never previously received antidepressant treatment, are assumed to initiate treatment at an annual rate 𝜓_𝑔_(𝑡), which is calculated as

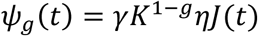

where 𝛾 is the annual rate of health seeking in women with depression symptoms; *K* is the relative rate of health seeking in males (𝑔 = 0); 𝜂 is the average probability of initiating antidepressants if seeking treatment, in the period up to 2008; and 𝐽(𝑡) is the relative rate of antidepressant prescription in year 𝑡, relative to the pre-2008 average (because we have no antidepressant data before 2008, we assume for simplicity that antidepressant initiation rates prior to 2008 were constant). The rate of initiating antidepressant treatment is assumed to increase by a factor 𝑃 if an individual has been previously treated. The rate of depression resolution is assumed to increase by a factor 𝐹 if an individual is currently taking antidepressants, and if an individual remains on antidepressants after recovery, the recurrence rate is assumed to reduce by proportion 𝜀. Due to high rates of antidepressant discontinuation during the first few months after treatment initiation [55–59, 65], a higher rate of discontinuation is assumed in the acute phase of depression (𝛿_0_) than in the recovery phase (𝛿_1_). Parameter values are summarized in Table 1 and explained more fully in section 1.3 of the supplementary materials.

### Calibration to depression prevalence data

The model is calibrated to align with nine nationally representative household survey estimates of depression prevalence. The South African Stress and Health (SASH) study, conducted over 2002-2004, used the Composite International Diagnostic Interview (CIDI) to estimate the cumulative incidence of depression over the last 12 months [29]. The National Income Dynamics Study (NIDS) was a panel study conducted over five waves (in 2008, 2010-11, 2012, 2014-15 and 2017), and used the Centre for Epidemiological Studies Depression (CES-D) 10-item Scale [31, 66], with a value of 10 or more to identify ‘probable depression’. The South African Human Development Pulse Survey was conducted in 2021 [30], and was repeated in 2022 and 2024, using the Patient Health Questionnaire-9 (PHQ-9), again with a value of 10 or more identifying ‘probable depression’.

The survey data were used to define two likelihood components. Firstly, the cross-sectional data from each survey were used to calculate prevalence stratified by sex and age group (15-24, 25-34, 35-44, 45-59 and 60+), and a likelihood statistic was defined to measure the agreement between the model-estimated depression prevalence (for the corresponding year, sex and age group) and the survey data. To ensure comparability between the model and survey estimates we made assumptions about the ratio of the prevalence we would expect to measure when using the survey tool to the true prevalence of depression (the 𝑅_1_*-* 𝑅_3_parameters in Table 1), taking into account that (a) 12-month prevalence should be higher than point prevalence, and (b) screening tools such as PHQ-9 and CES-D tend to over-estimate the true prevalence of depression [32]. Further detail is provided in section 1.4.2 of the supplementary materials.

The second likelihood component drew on the longitudinal nature of the NIDS data, and aimed to assess the extent to which depression persists/recurs in the same individuals. For each of the first four NIDS surveys, and for males and females separately, we calculated the odds ratio for the association between depression at the time of the survey and depression at the next survey. Initial exploration of the NIDS data suggested higher odds ratios when stricter definitions of depression are used (i.e. higher CES-D-10 thresholds), implying a negative relationship between depression prevalence and the odds ratio (parameter 𝛽 in Table 1). We adjusted for this when comparing the model and survey estimates of the odds ratios and calculating corresponding likelihood statistics; a more detailed explanation is provided in section 1.4.3 of the supplementary materials.

### Calibration to antidepressant use data

To ensure model alignment with local antidepressant use data, PubMed and Google were searched for studies on the prevalence of antidepressant use in South Africa. Two types of data were identified: surveys and cohort studies reporting prevalence of antidepressant use (18 data points from 7 studies [52, 53, 67–71]), and reports on antidepressant drug dispensing (11 data points from 3 studies [33, 54, 72]). All the data related to the period 2008-2023.

For survey and clinical cohort data, we used a random effect model to calculate a beta-binomial likelihood, representing the consistency between the modelled and survey estimates of antidepressant use. Sixteen of the 18 data points were from cohorts of patients receiving treatment for other chronic conditions (groups likely to have higher antidepressant use), and substantial heterogeneity in survey estimates was observed across studies. To address this, we controlled for differences between the modelled South African population and each survey population in treatment for other chronic conditions, public/private healthcare use and sex. Although the first two of these control variables are not used to stratify the modelled population, we estimated the proportions of the population in each category using data from the nationally representative General Household Surveys over the 2009-2015 period [73], and estimated levels of antidepressant use in each category based on the overall modelled prevalence of antidepressant use (by age and year) and assumed relative rates of antidepressant use by chronic disease treatment status and health sector (Table 1). Despite these adjustments, there was still substantial heterogeneity in the survey estimates of antidepressant use that could not be explained by the model, and we therefore included a random effect term in the likelihood calculation, to represent residual variation (the coefficient of variation for this random effect term is estimated in the model calibration process). A more detailed explanation is provided in section 1.5.2 of the supplementary materials.

For drug dispensing data, we estimated lower and upper bounds on the proportion of adults likely to be on antidepressants, for each data point, taking into account known sources of bias in the drug dispensing data. These were used to define the mean and standard deviation of a normal likelihood, to assess the consistency between the model and the data. (Using an approach similar to that described in the previous paragraph, the model estimates were also adjusted to control for differences in drug use between the public and private sectors.) A more detailed description is provided in section 1.5.3 of the supplementary materials.

### Model analyses

We adopted a Bayesian approach in model fitting and uncertainty analysis. Prior distributions were specified to represent ranges of uncertainty around key input parameters, based on reviews of relevant literature (Table 1). Likelihood functions were calculated to represent the consistency with the calibration data, as described previously. Posterior distributions, representing the synthesis of the priors and likelihoods, were estimated numerically, using Incremental Mixture Importance Sampling [74]. A sample of 1000 parameter combinations was drawn from the posterior distributions, and was used to generate the means and 95% confidence intervals in all analyses (see sections 1.4.4 and 1.5.4 of the supplementary materials).

A number of sensitivity analyses were performed:

- Model A: This is the ‘base’ model described previously. 7% of adults are assumed to be in the high-risk group [39].
- Model B: This is the same as Model A, except that we allow for a time trend in depression incidence, to represent the potential effect of changes in risk factors that are not explicitly represented in the model. A relative rate of depression incidence in 2025 (relative to 2000, after controlling for differences in age, sex and other modelled risk factors) is specified, and incidence is assumed to change linearly between 2000 and 2025. A prior distribution with mean 1 and standard deviation 0.2 is used to represent the uncertainty in this relative rate.
- Model C: This is the same as Model A, except that we omit the effect of COVID-19 (i.e. setting 𝜃_𝐷_ to zero).
- Model D: This is similar to Model A, but to test the hypothesis of extreme heterogeneity in depression incidence and a relatively low lifetime risk of depression, we set the proportion of adults in the high-risk group to 15% (towards the upper limit from international surveys [14, 15]) and set the relative rate of depression incidence in the low-risk group (𝛺) to zero. To avoid division-by-zero errors in the first equation, the priors on the incidence rates (𝜆_0_ and 𝜆_1_) are instead applied to the high-risk group and the previously-specified prior means are used as lower bounds for the revised priors.
- Model E: This is intermediate between models A and D. The same high-risk proportion is assumed in as in Model D (15%), but we allow for a non-zero depression incidence in the low-risk group, with a more conservative prior on the ratio of low- to high-risk depression incidence (mean 0.15, standard deviation 0.1).

The same calibration procedure is followed for each model. The plausibility of each of Models B-E, relative to Model A, is assessed using Bayes factors [75]. The models are also compared in terms of the estimated efficiency of antidepressant treatment, which is calculated by running a counterfactual in which there is no antidepressant treatment and estimating the number needed to treat (*N*𝑇) in order to prevent one case of depression:

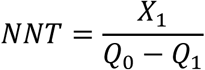

where 𝑋_1_is the number of people on antidepressant treatment mid-2024, 𝑄_1_ is the number living with depression mid-2024, and 𝑄_0_is the number living with depression mid-2024 in the counterfactual scenario.

## Results

### Model calibration

The model fit to the calibration datasets is summarized in Figure 2. The model fits the overall depression prevalence data in the surveys reasonably well, although the 2010 survey measured a significantly lower prevalence than the model estimated (Figure 2a); age- and sex-specific model comparisons against the survey data are included in Figure S7. Model estimates of the prevalence of antidepressant use were consistent with the three studies that provided information on antidepressant drug volumes (Figure 2b), but there was a high degree of heterogeneity in survey/cohort estimates of antidepressant use, with most of these estimates being higher than the model estimates because they sampled patients with chronic conditions. Model estimates of the association between initial and subsequent depression (over two-year intervals) were in good agreement with longitudinal survey estimates, which showed relative weak associations (Figure 2c). Posterior model estimates of model parameters were mostly consistent with prior distributions (Table S9).

**Figure 2:**
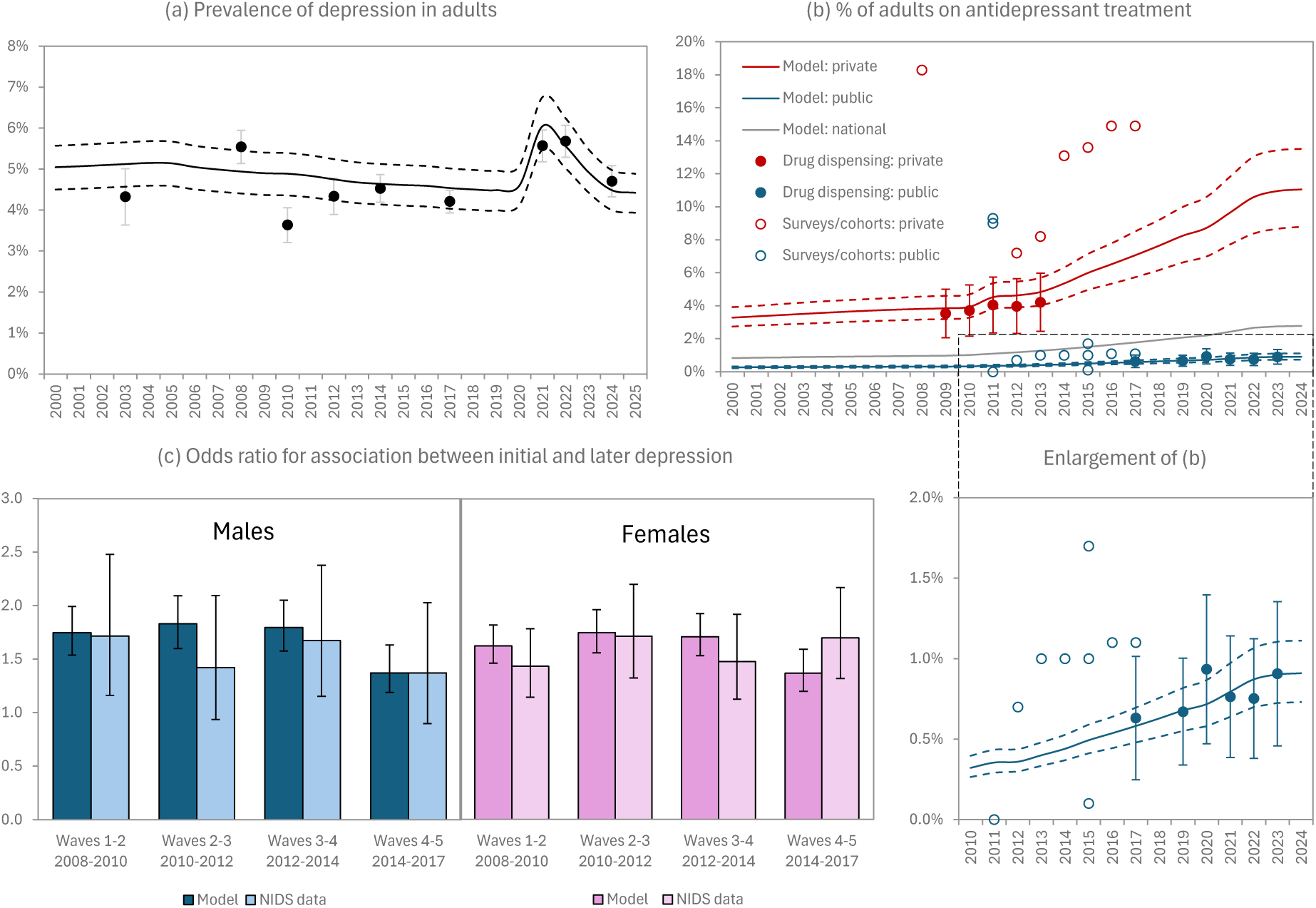
Model calibration. In panels a and b, solid lines represent posterior means of model estimates, dashed lines represent posterior 95% confidence intervals, and dots represent calibration data. In all panels, error bars represent 95% confidence intervals. In panel a, survey estimates have been adjusted downward by the posterior estimates of the correction factors (see Table S9). The bottom right panel expands the public sector data and estimates from panel b to improve readability. NIDS = National Income Dynamics Study.

### Key indicators

The model estimates that the prevalence of depression in South Africans aged 15 and older decreased slightly from 2002 (5.1%, 95% CI: 4.5-5.6%) to 2024 (4.5%, 95% CI: 4.0-5.0%), although with a transient rise in depression prevalence during the COVID-19 period (Figure 2a). In 2024, depression prevalence was higher in women (5.3%, 95% CI: 4.7-5.9%) than in men (3.6%, 95% CI: 3.2-4.0%), and depression prevalence was also substantially higher in older adults than in younger adults (Figure 3). Although depression prevalence in people living with HIV was higher than that in the total population in the early stages of the ART rollout (e.g. 7.1% versus 4.9% in 2010), the difference narrowed as ART coverage increased (5.9% versus 4.5% in 2024). Because initial depression was only weakly associated with subsequent depression (Figure 2c), cumulative depression risk could not be confined to the high-risk group, and a substantial cumulative risk was estimated for low-risk individuals too: in 2024 an estimated 70.6% of South African adults (95% CI: 67.8-73.6%) had ever experienced depression. The lifetime prevalence was lower in adults aged 15-39 (55.5%, 95% CI: 52.1-59.2%).

**Figure 3:**
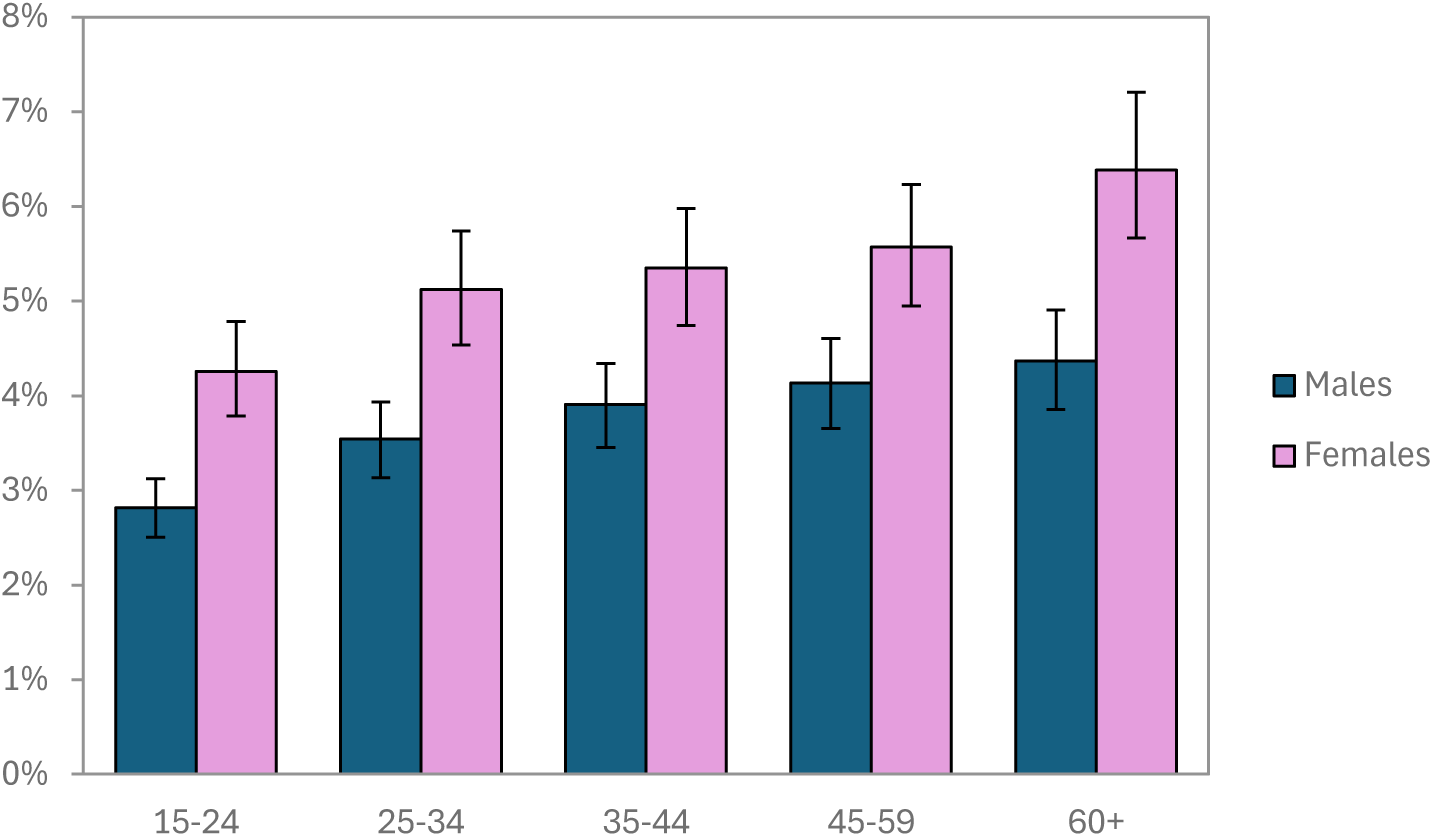
Depression prevalence by age and sex, in 2024. Error bars represent 95% confidence intervals.

Due to the lack of data on antidepressant use before 2008, it is difficult to estimate long-term trends. Nevertheless, the model suggests that the proportion of adults using antidepressants increased from 1.0% (95% CI: 0.8-1.2%) in 2008 to 2.8% (95% CI: 2.2-3.4%) in 2024. In 2024, the proportion of women on antidepressants (4.4%, 95% CI: 3.3-5.6%) was more than four times the proportion in men (1.0%, 95% CI: 0.4-1.8%). The proportion of medical scheme beneficiaries (private sector) on antidepressants in 2024 was 11.0% (95% CI: 8.8-13.5%), compared to only 0.9% (95% CI: 0.7-1.1%) in the rest of the population. The number of adults initiating antidepressants in 2024 was 443 000 (95% CI: 266 000-722 000), compared to a total incidence of 3.84 million new adult episodes of depression in 2024 (95% CI: 3.33-4.52 million), a ratio of 0.12 new treatment initiations per incident case.

### Sensitivity analyses

Model results were similar when the model was refitted to allow for a linear trend in depression incidence unrelated to the modelled risk factors (Model B), to exclude the effect of the COVID-19 pandemic (Model C), or to allow for a larger high-risk group (Model E), as shown in Table 2 and Table S9. However, the Bayes factor comparing Models B and C to Model A were 0.08 and 0.13 respectively, indicating “positive” evidence [75] that Models B and C were less consistent with the data than Model A. Results differed substantially when depression incidence was assumed to occur only in a ‘high-risk’ group comprising 15% of the population (Model D). In this specification, the model failed to converge when calibrated simultaneously to both depression prevalence data and longitudinal data on associations between initial and later depression, because the model was not able to reproduce the latter. The model was therefore calibrated only to prevalence data, and the poor correspondence with the longitudinal data is shown in Figure S8. In Model D, the estimated lifetime prevalence of depression in 2024 was 14.1% (because depression was assumed to occur only in the 15% of the population that comprised the high-risk group), and the NNT was 4.5, compared to around 11 in the other models. Posterior estimates of all parameters are compared across models in Table S9; although treatment parameters were similar across models, the incidence and duration parameters were markedly different in Model D.

**Table 2:** Comparison of key indicators across models.

|  | Model A:<br>Main<br>model | Model B:<br>Add time<br>trend | Model C:<br>No COVID<br>effect | Model D:<br>No low-risk<br>depression | Model E:<br>Larger<br>high-risk |
| --- | --- | --- | --- | --- | --- |
| Prevalence of depression: 2002 | 5.1%<br>(4.5-5.6) | 5.2%<br>(4.7-5.8) | 5.1%<br>(4.6-5.7) | 5.6%<br>(4.8-6.5) | 5.1%<br>(4.6-5.7) |
| Prevalence of depression: 2024 | 4.5%<br>(4.0-5.0) | 4.4%<br>(3.9-5.0) | 4.4%<br>(4.1-5.0) | 5.0%<br>(4.4-5.8) | 4.5%<br>(4.0-5.0) |
| Lifetime prevalence of depression: 2024 | 70.6%<br>(67.8-73.6) | 70.6%<br>(67.7-73.6) | 70.4%<br>(67.2-73.4) | 14.1%<br>(13.7-14.6) | 70.4%<br>(67.5-73) |
| Bayes factor (relative to Model A)* | - | 0.08 | 0.13 | - | 1.03 |
| % of adults using antidepressants: 2024 | 2.8%<br>(2.2-3.4) | 2.8%<br>(2.2-3.3) | 2.6%<br>(2.0-3.2) | 2.6%<br>(2.0-3.2) | 2.7%<br>(2.1-3.4) |
| Number needed to treat to prevent one<br>case of depression: 2024 | 10.9<br>(8.9-12.7) | 11.9<br>(9.9-13.9) | 11.6<br>(9.5-13.7) | 4.5<br>(4.2-4.8) | 11.7<br>(9.2-14.1) |

\* The Bayes factor represents the ratio of the integrated likelihood in one model to that in another (reference) model; a value below 1 indicates a lower likelihood when compared to the reference model. 95% confidence intervals are shown in brackets.

## Discussion

Based on a synthesis of South African data sources, we estimate that nearly 5% of South African adults currently experience depression, with depression prevalence being substantially higher in older adults and women. Depression prevalence is estimated to have declined slightly since 2002, when South Africa’s first national household survey of depression prevalence was conducted, although there is some evidence of a transient increase during the period of the COVID-19 pandemic. Despite a trend towards rising antidepressant use, the ratio of antidepressant initiations to new episodes of depression in 2024 was only 0.12. There are substantial inequalities in access to and use of antidepressants, with people in the private sector being more than 10 times as likely to receive treatment as those in the public sector, and women being more than 4 times as likely to receive treatment as men.

Our finding of a substantial sex difference in depression prevalence is consistent with international surveys [15], but age patterns are less consistent across countries [15]. Our model estimate of a slight decline in depression prevalence over time is likely to be due to an assumed decline in the prevalence of two key depression risk factors (violence exposure [76, 77] and physical inactivity [78]), as well as the previously noted increase in the use of antidepressants. The model estimate of trend in prevalence was robust to alternative model specifications, including when allowing for a non-zero trend in depression incidence (unrelated to the modelled risk factors). Although we have not presented age-standardized measures of depression prevalence, age-standardization would lead to a more significant decline, given that South Africa’s adult population has aged over the 2002-2024 period, and depression prevalence is higher in older adults. A strength of the model is that it integrates a number of the known social determinants of depression identified in South Africa’s National Mental Health Policy Framework and Strategic Plan [28], and is capable of evaluating policies to address these risk factors. However, a number of social determinants are not modelled, most significantly unemployment and poverty. A further limitation is that we do not attempt to simulate the specific mechanisms by which the COVID-19 pandemic increased depression prevalence (e.g. increased isolation, bereavement, economic stress, or symptoms of ‘long COVID’ [48, 79]).

Our estimates of rising antidepressant use are consistent with international trends [9–11]. Although South African data are too limited to identify the most likely explanation, the increase may be due to rising public awareness of mental health and reduced stigma around seeking treatment [80], increasing use of selective serotonin reuptake inhibitors (which are better tolerated) [59], or evolving guidelines on antidepressant use (which increasingly recommend longer treatment durations). The lower use of antidepressants in men is partly due to the lower prevalence of depression in men, but is also significantly driven by lower rates of health seeking in men who experience depression symptoms [51], a pattern common to other diseases in the African context and globally [81, 82]. Further efforts are needed to de-stigmatize mental health seeking in men [83], and to address the inequitable gender norms that underpin this [84, 85].

Although levels of antidepressant use in South African private healthcare are similar to those in high-income countries [7–9], only 16% of South African adults have private health insurance [86]. Levels of antidepressant use in the uninsured population are very low, despite a substantially greater prevalence of depression in people of lower socioeconomic status [30, 87–89]. This points to a substantial gap in addressing mental healthcare needs, consistent with previous South African studies, which have found very poor access to mental health care in the uninsured population [53, 90, 91]. In South Africa, antidepressants are classified as scheduled substances requiring both medical diagnosis and management, but also enhanced control of supply such as record-keeping (registers), limitations on re-prescribing (6-monthly review by a specialist psychiatrist) and dispensing [92], which prevents nurses from prescribing the drugs. This is a major challenge, given that most public clinics are nurse-led with limited access to doctor support. Access to psychiatrists is also limited, with only 1.5 psychiatrists per 100 000 population, of whom only 20% work in the public sector [93]. The National Mental Health Policy Framework and Strategic Plan calls for regulatory review with a view towards adoption of nurse-initiated antidepressant treatment [28].

Our estimate of the prevalence of depression in South Africa is similar to that measured in the only national household survey that used a diagnostic interview [29], but slightly higher than that published by the Global Burden of Disease (GBD) study (3.1% [95% CI: 2.7-3.5%] in 2019) [2]. This may be because our estimate of prevalence relates only to ages 15 and older, while young children tend to have a lower prevalence [32, 48]. It may also be a reflection of differences in data sources, as we rely only on South African data, while the GBD approach relies on regression modelling, pooling data from other countries with similar characteristics. A strength of our analytic approach is that it relies on dynamic modelling, thus incorporating effects of treatment on depression and allowing for a greater range of data in calibration, including data on antidepressant use, and allowing for more explicit representation of disease heterogeneity.

Despite our estimates of the current prevalence of depression in South Africa being roughly consistent with other sources, our estimate of the lifetime prevalence of depression (70.6%) is substantially higher than the 10% that is often quoted for South Africa [29] and the 10-15% range from international surveys [14, 15]. This finding is driven largely by the NIDS, the only nationally-representative longitudinal study providing data on the stability of depression symptoms. We found that if we considered a model where only 15% of the population was ever at risk of depression (Model D), it failed to reproduce the observed longitudinal associations between initial and subsequent depression episodes, overestimating recurrence relative to the NIDS data. Model D is also not consistent with longitudinal studies in adolescents, in which those considered ‘high risk’ based on persistently high depression symptoms account for less than half of depression diagnoses [94, 95]. A more plausible explanation for the low lifetime prevalence measured in cross-sectional surveys is that these surveys are subject to substantial recall bias, as has been shown previously in various international settings [17–21]. Even in patients hospitalized for depression, only half who are re-interviewed correctly recall the episode [96], and levels of recall are likely to be substantially worse when depression is less severe or remains untreated [97, 98], or interviewees are less literate [99]. Prospective studies have confirmed this recall bias [17, 20, 100], but even prospective studies are likely to under-estimate the true lifetime risk because follow-up is typically short, or there are long intervals between follow-up rounds in which people may forget depression episodes. If one assumes a 50% under-reporting due to recall bias, our estimate of a lifetime prevalence of 55% in young adults (ages 15-39) appears compatible with a retrospective survey of young adults in the Eastern Cape province of South Africa, in which a 31% lifetime prevalence was reported [101].

The stability or instability of depression risk has important implications for strategies to reduce the incidence of depression. If the incidence of depression were truly concentrated in a small sub-population that experiences high rates of recurrence, then it would perhaps be most cost-effective to focus on diagnosing and treating depression in that sub-population, to prevent recurrence (as suggested by the lower NNT for Model D). Conversely, if a larger proportion of the population is at risk, broader preventive strategies (e.g. focusing on social determinants of depression) may yield greater population-level benefit.

Our study has several limitations. Our estimates of weak associations between initial and later depression are informed by NIDS data, which used symptom screens rather than diagnostic interviews, and although we adjust for the biases introduced by this imprecision, uncertainty remains. We lack nationally representative data on antidepressant use, and the limited data are all from the period 2008-2023. Most of the antidepressant survey data are from people already receiving medication for other chronic conditions, who are unlikely to be representative of the general population, and although we account for this bias in our modelling, it nevertheless introduces some uncertainty. We do not differentiate depression according to level of severity or co-occurrence with other psychiatric disorders. We have not attempted to model cognitive behavioural therapy (CBT) or other evidence-based psychological treatments here, due to lack of data on access. CBT can be as effective, if not more effective, than antidepressants, both in treating depression [102, 103] and preventing recurrences [5, 104]. This is especially pertinent given concerns around the possible ‘over-medicalization’ of social and emotional challenges [9, 80]. CBT should therefore be considered together with antidepressants when evaluating the impact and cost-effectiveness of different strategies to reduce the incidence of depression in South Africa.

This study lays a foundation for future modelling of the potential impact and cost-effectiveness of strategies to reduce the prevalence of depression in South Africa. It also provides an example for other low- and middle-income countries of how system dynamics modelling can be used to collate diverse data sources. In these countries, routine mental health information systems are often limited [105], necessitating reliance on infrequent surveys. While routine mental health data systems can and should be strengthened [105], models can complement these efforts by providing baseline estimates of depression disease burden, and by quantifying how much of this burden remains untreated. System dynamics models have been widely applied in the context of infectious diseases, and have been instrumental in advocating for greater funding for diseases such as HIV [106], but they remain under-utilized tools for advocacy and strategic planning in mental health.

## Data Availability

All data produced in the present study are available upon reasonable request to the authors.

https://saldru.uct.ac.za/surveys/national-income-dynamics-study-nids

## Acknowledgements

We are grateful to Dan Stein and Naomi Folb, who both shared unpublished data and gave helpful feedback on earlier drafts of this paper. We thank Ian Neethling for sharing data on physical activity levels.

