## Supplementary materials for "Patterns of depression prevalence and antidepressant use in South Africa, 2002-2024: a system dynamics modelling perspective"

### 1. Model description

The main text describes the model structure. Here we provide more details on the estimation of the model parameters and the calibration process,

#### 1.1 Depression incidence

We distinguish here between innate factors that drive heterogeneity in susceptibility to depression and external factors (factors that can change over time in the same individual, as their life circumstances change). The first section discusses the modelling of heterogeneity in innate susceptibility, and the following sections describe the modelling of each of the external factors.

##### 1.1.1 Heterogeneity in innate susceptibility to depression

We divide our population into two broad groups: high-risk and low-risk. Although we do not attempt to define the high-risk group in exact terms, this can be thought of as the subset of the population that is at high risk of depression due to genetic or neurological factors, or stressful early-life experiences (potentially interacting with genetic factors [1]). This distinction between high-risk and low-risk groups is motivated by the modelling of Demic and Cheng [2], who found that it was only possible to match a model to observed distributions of depression episode counts when allowing for these two risk groups. Many people experience a single depression episode and never experience a recurrence [3], while others experience frequent recurrences [4], and in these individuals depression can be considered chronic; this corresponds to our low-risk/high-risk distinction. In line with Demic and Cheng, we assume that 7% of the population is in the high-risk group, and that this proportion is constant with respect to age and sex, and not varying over time. This is also consistent with a systematic review of long-term trajectories of depressive symptoms, which found that "a small proportion (<10%) of individuals across different populations and age groups persistently report chronic, high levels of depressive symptoms for extended periods of time" [4].

*Incidence in low-risk adults*

Few population-based estimates of depression incidence have been published. In one cohort of South African women, 18% of those who did not have depression at baseline developed depression within a year [5]. This is likely to be an over-estimate of the true incidence of depression, as depression was diagnosed using the CES-D, which is known to yield a high rate of false positives [6]. Another South African study estimated a 6-month depression incidence of 8.1% using the more accurate MINI diagnosis tool [7] (equivalent to an annual incidence of 15.5%), but this is likely to be an over-estimate because the cohort comprised people who had recently been diagnosed HIV-positive, which is a major risk factor for depression (see section 1.1.5). In an international systematic review, only four studies were found that had estimated the incidence of depression longitudinally, with annual incidence rates varying between 1.6% and 7.1% [8]; the average annual incidence rate was 2.7% in males and 3.4% in females. These might be over-estimates of the depression incidence rate in the low-risk group (since the overall depression incidence is a weighted average of the incidence rates in the low- and high-risk groups). On the other hand, they could also be under-estimates of the true depression incidence rates because in most of the four studies there were relatively long intervals between interviews (e.g. 18-60 months [9]), which implies potential recall bias. We represent the uncertainty around the annual incidence rates in the 'base group' (low-risk individuals with no risk factors) using gamma distributions with means of 0.027 and 0.034 in males and females respectively, and with standard deviations of 0.010 and 0.013 respectively (implying a coefficient of variation of 0.38). The female distribution has 2.5 and 97.5 percentiles of 0.013 and 0.064 respectively, roughly consistent with the range of female estimates in the cited review (1.5-7.1%). Due to the relative lack of male data, we use the same coefficient of variation for men as in women when calculating the standard deviation for the male prior.

*Incidence in high-risk adults*

The ratio of depression incidence in the low-risk group to that in the high-risk group is highly uncertain. However, the ratio must be less than 1, since the low-risk group by definition has a lower incidence than the high-risk group. We would also expect the ratio to be substantially lower than 1, given the marked heterogeneity in incidence rates estimated in previous studies [2]. We therefore represent the uncertainty in this ratio using a beta distribution with a mean of 0.3 and a standard deviation of 0.2. This distribution has 2.5 and 97.5 percentiles of 0.02 and 0.75 respectively (corresponding to incidence rates in the high risk group that are 50 times and 1.33 times those in the low-risk group, respectively). The prior distribution is therefore sufficiently wide to represent anything from modest to extreme heterogeneity in depression incidence.

In Model D (described in the main text), we consider an alternative parameterization, in which the incidence rates in the low-risk group are set to zero, and we instead place priors on the incidence rates in the high-risk group. For the purpose of setting these priors, we consider the previously quoted annual incidence rates of 2.7% and 3.4% to be lower bounds (because the denominator is the total population, not the population in the high-risk group, and because of the potential recall bias described previously). Because depression episodes must be at least 8 weeks apart in order to be classified as distinct episodes [10], the upper bound on the annual incidence rate in the high-risk group is 6.5 (52/8). We therefore represent the uncertainty in the annual incidence rate in the high-risk group using a gamma distribution with a mean of 1.75 and standard deviation of 1.75 (there are separate priors for the male and female rates, but the parameters for the two priors are the same). The 2.5 and 97.5 percentiles of this distribution are 0.04 and 6.46 respectively. The lower limit is thus roughly consistent with the lower bound of 3% (based on the incidence rate in the general population) and the upper limit is consistent with the maximum incidence rate of 6.5 per annum.

##### 1.1.2 The effect of age

The effect of age on the prevalence of depression differs across settings: in high-income settings, depression tends to be more prevalent at younger ages than at older ages, while in resource-limited settings the effect of age is more heterogeneous [11]. The rate of *first* depression was found to be higher at younger ages than at older ages in a US cohort [3], although it is difficult to tease out how much of this is to be expected given the innate heterogeneity in lifetime risk noted previously. In older adults the relationship between age and incidence is less clear, with incidence possibly being higher (and recovery slower) with advancing age [12]. Younger age is associated with higher risk of recurrence in some studies [13].

South African studies are heterogeneous in the age effects that they have found. Most South African studies find a positive relationship [14-19], but one found a negative association in a sample of primary healthcare attenders [20], and in the nationally representative SASH survey a weak and non-linear relationship between age and depression was found, with depression risk being highest in middle-aged adults [21]. Although these studies generally suggest a positive relationship between age and depression in South Africa, it is unclear how much of this is attributable to other risk factors included in our model (e.g. the higher prevalence of non-communicable diseases at older ages).

In our model, we assume that the incidence of depression at age $x$ is proportional to

$$B^{(x-25)/10}$$

where $B$ is the factor by which incidence increases per 10-year increase in age ($x$). We represent the uncertainty in this parameter using a gamma prior with a mean of 1 and standard deviation of 0.1. A value of 1 corresponds to an assumption of no age effect (after controlling for the other sources of heterogeneity described below).

#### 1.1.3 Violence

Data from South Africa suggest strong effects of childhood abuse [17, 19, 22, 23], intimate partner violence [22] and more general exposure to community violence [18, 19, 24, 25] on depression. Randomized controlled trials (RCTs) of violence prevention programmes have also found reductions in symptoms of depression [26-29], lending support to the argument that exposure to violence increases the incidence of depression.

In our model we focus on the role of community violence, as this is the form of violence exposure that is most strongly associated with depression and it is also the most commonly reported violence exposure (based on data from the 2020-21 Quality of Life Survey, described in more detail in Appendix A). If $\lambda_{g,i}(x)$ is the annual incidence of depression in people in risk group $i$, of age $x$ and sex $g$, who have not experienced community violence in the last year, then the average incidence of depression in all people of risk group $i$, age $x$ and sex $g$ is

$$\lambda_{g,i}(x)\big(1 + V_g(x,t)[\theta_V - 1]\big)$$

where $V_g(x,t)$ is the proportion of people who have experienced community violence in the last year, in year $t$, and $\theta_V$ is the relative risk of depression in people who have experienced community violence in the last year. The $V_g(x, 2020)$ and $\theta_V$ parameters are estimated by fitting regression models to the data from the 2020-21 Quality of Life Survey, as described in Appendix A. Based on the results in Appendix A, we set the $\theta_V$ parameter to 1.56.

We estimate time trends in the prevalence of community violence based on South African Police Service statistics on reported numbers of assaults, per 100 000 population (Figure S1). The $V_g(x,t)$ variable is calculated by multiplying $V_g(x, 2020)$ by the ratio of the reported incidence rate of assault in year $t$ to that in 2020. Because we lack data for the period before

2004, the multipliers in the years before 2004 are assumed to be the same as those estimated for 2004. Similarly, the multiplier in the period after 2022 is assumed to be the same as that in 2022.

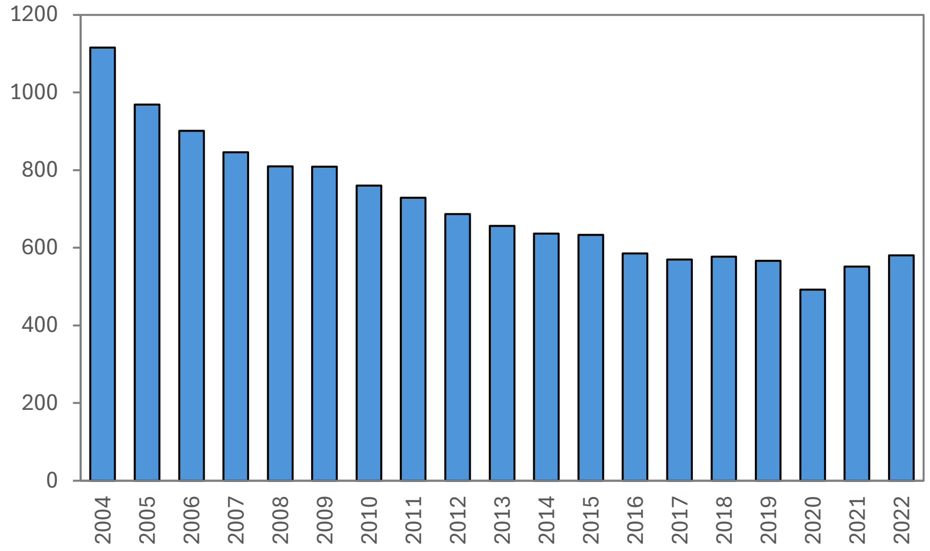

Figure S1: Reported rates of assault in South Africa, per 100 000 population
Source: SAPS website (https://www.saps.gov.za/)

#### 1.1.4 Alcohol

South African studies show a positive association between alcohol use and the prevalence of depression [21, 22, 30]. Although it is difficult to disentangle how much of this association is due to an effect of alcohol on depression versus an effect of depression on alcohol (or other confounding factors), longitudinal studies from high-income settings suggest that binge drinking strongly predicts future depression [3, 31, 32]. In addition, a meta-analysis of interventions that principally addressed excessive alcohol consumption among youth found that these interventions significantly reduced depression [33], which suggests an effect of alcohol on risk of depression. One South African RCT was included in this meta-analysis: a brief alcohol counselling intervention in young adults with hazardous alcohol use was found to significantly reduce depression [34], with an effect size similar to the average estimated in the meta-analysis. In this South African trial, the intervention reduced AUDIT (alcohol use) scores by 18% and reduced depression symptoms by 7% (95% CI: -1 to 14%). This suggests that for each 10% decrease in the level of alcohol consumption, there is a 4% reduction in the incidence of depression (range 0 to 8%).

Binge drinking is defined here as consuming at least 5 drinks on a single day in the last month, in line with our previous modelling [35]. If $\lambda_{g,i}(x)$ is the annual incidence of depression in people in risk group $i$, of age $x$ and sex $g$, who have a 10% prevalence of regular binge drinking, then the average incidence of depression in all people of risk group $i$, age $x$ and sex $g$ is

$$\lambda_{g,i}(x)\theta_A{}^{(0.1-A_g(x))/0.1}$$

where $A_g(x)$ is the proportion of people who engage in regular binge drinking (at least once a month), and $\theta_A$ is the relative risk of depression for each 10% reduction in the prevalence of binge drinking. For example, if we set $\theta_A$ = 0.96 (based on the previously described South African RCT) and assume the prevalence of binge drinking changes from 100% to 82% after an alcohol counselling intervention, the model predicts the relative rate of depression after the intervention compared to that pre-intervention is 0.93 ($0.96^{(0.1-0.82)/0.1}/0.96^{(0.1-1)/0.1}$). This is consistent with the 7% observed reduction in the incidence of depression observed in the cited trial.

The $A_g(x)$ parameters are obtained from the 2016 Demographic and Health Survey (DHS), and are derived by fitting logistic regression models to the proportions of people who report binge drinking in the last 7 days. The models include cubic polynomials to represent age differences, and are fitted separately for males and females (Table S1).

Table S1: Parameters for relationship between age and odds of binge drinking

| | Males | Females |
|---|---|---|
| Regression model output | | |
| Age | 1.42 (1.31-1.54) | 1.26 (1.08-1.47) |
| Age$^2$ | 0.9929 (0.9911-0.9948) | 0.9948 (0.9911-0.9985) |
| Age$^3$ | 1.00004 (1.00003-1.00005) | 1.00003 (1.00000-1.00006) |
| Constant | 0.0022 (0.0008-0.0063) | 0.0027 (0.0004-0.0189) |
| Under-reporting adjustment | 4.02 | 12.28 |

The results of these regression models understate the true prevalence of binge drinking. This is partly because the survey asked about drinking in the last week, but many South Africans engage in binge drinking on a less frequent basis. In addition, there is social desirability bias, which leads to under-reporting of both the frequency of drinking and the amount of alcohol consumed [36, 37]. This social desirability bias may be particularly substantial in women [38, 39]. Survey estimates of alcohol consumption may also be biased due to non-response, with heavy drinkers being less likely to be sampled/interviewed [40]. To address the under-reporting issues, we apply an odds ratio to the survey-estimated binge-drinking proportions to bring the aggregated model estimates in line with previous estimates of the binge drinking prevalence (54% in men and 35% in women aged 15-49 in 2021 [35]). These previous estimates for 2021 are considered more reliable as they are based on calibrating a national model to alcohol sales data. The assumed odds ratios are 4.0 in males and 12.3 in females; Figure S2 shows the implied prevalence of binge drinking by age, before and after adjusting for under-reporting. The adjusted prevalence levels are reasonably consistent with a national survey of school-going youth in 2008, in which 33.5% of males and 23.7% of females reported binge drinking in the last month [41] (the results are less likely to be affected by social desirability because the questionnaires were completed anonymously, not through face-to-face interviews). As our previous analysis does not suggest any significant change in drinking patterns over time [35], we have not accounted for possible temporal changes in drinking levels.

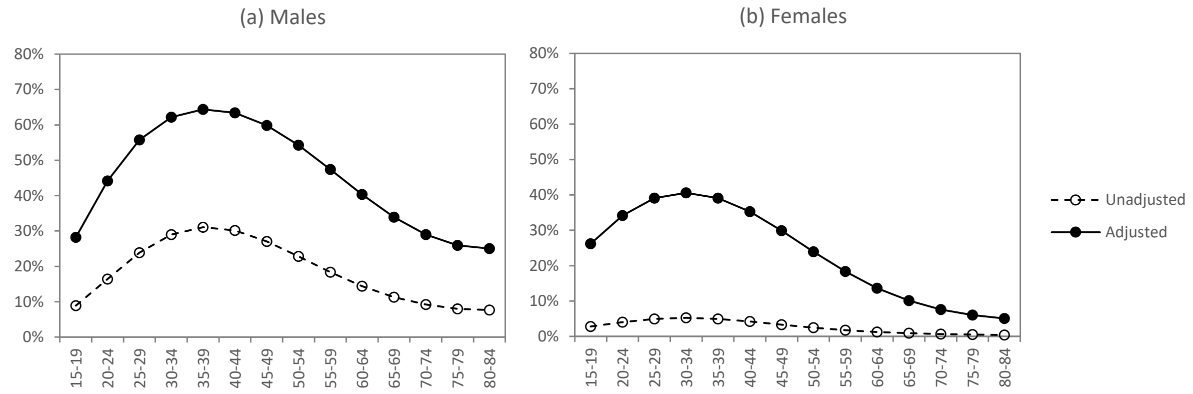

Figure S2: Proportions of people engaging in binge drinking at least once a month
Unadjusted estimates are based on a regression model fitted to 2016 DHS data. Adjusted estimates are derived by scaling up the unadjusted estimates to match overall binge drinking prevalence levels estimated in a previous modelling study, using alcohol sales data [35].

##### 1.1.5 HIV

In an early meta-analysis by Ciesla and Roberts [42], based on data from the period before the availability of triple-drug antiretroviral treatment (ART), HIV was found to be strongly associated with diagnosed depression (OR 1.99, 95% CI: 1.32-3.00). However, in a recent review and meta-analysis of South African studies, we found HIV to be only weakly associated with depression (OR 1.24, 95% CI: 0.41-2.87) [43]. Both meta-analyses were based on pooling unadjusted odds ratios, and it is possible that associations between HIV and depression might be stronger or weaker when controlling for other variables. In addition, our South African meta-analysis found substantial heterogeneity across the five studies. Two South African studies were excluded from the review because they relied on self-reported knowledge of HIV status; both found positive associations between depression and HIV [44, 45], and in both studies relatively few people with diagnosed HIV appeared to be on ART. African studies suggest that in people living with HIV, symptoms of depression are greatest in people with HIV symptoms [7, 30, 46], low CD4 counts [46, 47] and recent HIV diagnosis [7, 48-51]. Longitudinal studies suggest that ART initiation leads to substantially reduced incidence of depression [46, 52, 53], while poor ART adherence and missed clinic visits are associated with increased depression risk [54]. In a meta-analysis of studies of HIV stigma in people living with HIV, poor mental health was consistently associated with stigma [55], suggesting that shame and fear associated with knowledge of HIV status are major mediators of the relationship between HIV and depression. In contrast, ART initiation is strongly associated with lower HIV stigma [56], and reductions in depression after starting ART are likely to be mediated both by greater acceptance of one's HIV status and ART-related CD4 recovery and improved physical health.

In our model we assume that people living with HIV are at an elevated risk of developing depression, but account for heterogeneity in HIV effects over the course of disease. The variable $H_1$ represents the relative risk of depression in people who are diagnosed and untreated (relative to HIV-negative individuals) and $H_2$ represents the relative risk of depression in people who are either on ART or undiagnosed (again, relative to HIV-negative individuals). Relatively little is known about depression in people with undiagnosed HIV, but one might expect a modest increase in depression risk due to HIV-related symptoms. One South African study found depression symptoms were associated with undiagnosed HIV [57], although the authors interpreted this as an effect of depression on HIV care seeking rather than an effect of undiagnosed HIV on depression. In the interests of model simplicity, we use the same $H_2$ parameter to represent the increased depression risk in undiagnosed HIV and when on ART.

We use network meta-analysis to estimate the $H_1$ and $H_2$ parameters [58]. We identified three types of comparison for the purpose of this meta-analysis: (1) studies comparing people with diagnosed but untreated HIV to HIV-negative individuals (from the early meta-analysis of pre-ART studies [42]); (2) studies comparing people on ART to HIV-negative individuals (from South African data sources in our meta-analysis [43], limiting to studies in which almost all people living with HIV were known to be on ART); and (3) studies comparing people before and after starting ART. The data from these different sources is summarized in Table S2 below. Network meta-analysis was performed in STATA 17, using the *network* package [58].

Table S2: Studies used to estimate the effect of HIV diagnosis and ART on depression

| Comparison | Study | OR | Log OR | SE (log OR) |
|---|---|---|---|---|
| Diagnosed but untreated vs HIV-negative | Atkinson 1988* | 4.00 | 1.386 | 1.121 |
| | Williams 1991* | 1.13 | 0.126 | 0.745 |
| | Chuang 1992* | 0.99 | -0.006 | 0.801 |
| | Rosenberger 1993* | 1.65 | 0.504 | 0.775 |
| | Lipsitz 1994* | 1.41 | 0.341 | 0.342 |
| | Maj 1994* | 3.81 | 1.337 | 0.414 |
| | Perkins 1994* | 3.06 | 1.119 | 0.806 |
| | Rabkin 1997* | 0.91 | -0.093 | 0.483 |
| | Kelley 1998* | 2.20 | 0.789 | 0.474 |
| Treated vs HIV-negative | Jelsma 2005 [53] | 0.53 | -0.635 | 0.419 |
| | Tlali 2025 [59] | 2.03 | 0.708 | 0.301 |
| | Petersen 2019 [60] | 2.09 | 0.737 | 0.159 |
| | Medscheme data [43] | 1.05 | 0.049 | 0.014 |
| Treated vs diagnosed but untreated | Wagner 2012 [46] | 0.51 | -0.673 | 0.280 |
| | Gutierrez 2014 [52] | 0.53 | -0.635 | 0.322 |
| | Jelsma 2005 [53] | 0.37 | -0.994 | 0.414 |

* From the meta-analysis by Ciesla & Roberts [42]. OR = odds ratio.

The network meta-analysis estimates that the average OR comparing depression prevalence in diagnosed untreated HIV to that in HIV-negative individuals is 2.04 (95% CI: 1.36-3.08), and the average OR comparing depression prevalence in people on ART to that in HIV-negative individuals is 1.19 (95% CI: 0.81-1.75). There was no significant evidence of network inconsistency (p = 0.29). These results are similar to the odds ratios in the previously cited meta-analyses, 1.99 [42] and 1.24 [43] respectively (the latter being based mostly on data from ART patients). We therefore set the $H_1$ and $H_2$ parameters to 2.0 and 1.2 respectively.

##### 1.1.6 Other chronic conditions

In an analysis of World Health Survey data from 60 countries, Moussavi *et al* found the prevalence of depression in people with asthma, ischaemic heart disease (IHD), arthritis or diabetes was around 3-6 times that in people without any of these other chronic conditions [61], with diabetes being the condition most weakly associated with depression. This is consistent with South African studies that have found strong associations between these conditions and depression [43]. However, South African studies have found only weak association between depression and hypertension [14, 21, 60, 62], which is probably because most hypertension is asymptomatic, causing relatively little impairment. Our review of South African data also found that diabetes was not as strongly associated with depression as asthma, chronic obstructive pulmonary disease, arthritis, stroke and IHD [43].

Based on this evidence, we assume that the incidence of depression is increased in proportion to the fraction of people who have asthma, arthritis, IHD or past stroke. If $\lambda_{g,i}(x)$ is the annual incidence of depression in people in risk group $i$, of age $x$ and sex $g$, who have none of the listed chronic conditions, then the average incidence of depression in all people of risk group $i$, age $x$ and sex $g$ is

$$\lambda_{g,i}(x)\big(1 + C_g(x,t)[\theta_C - 1]\big)$$

where $C_g(x,t)$ is the proportion of people who have asthma, arthritis, IHD or past stroke in year $t$, and $\theta_C$ is the relative risk of depression in people who have one of the four conditions. The $C_g(x,t)$ parameters are estimated by fitting a logistic regression model to self-reported data from the 2013 General Household Survey (Figure S3). For simplicity, we assume that the age- and sex-specific prevalence levels remain constant over time, as the questions asked in the GHS have not been stable enough to permit a thorough assessment of changes in the prevalence of these conditions over time.

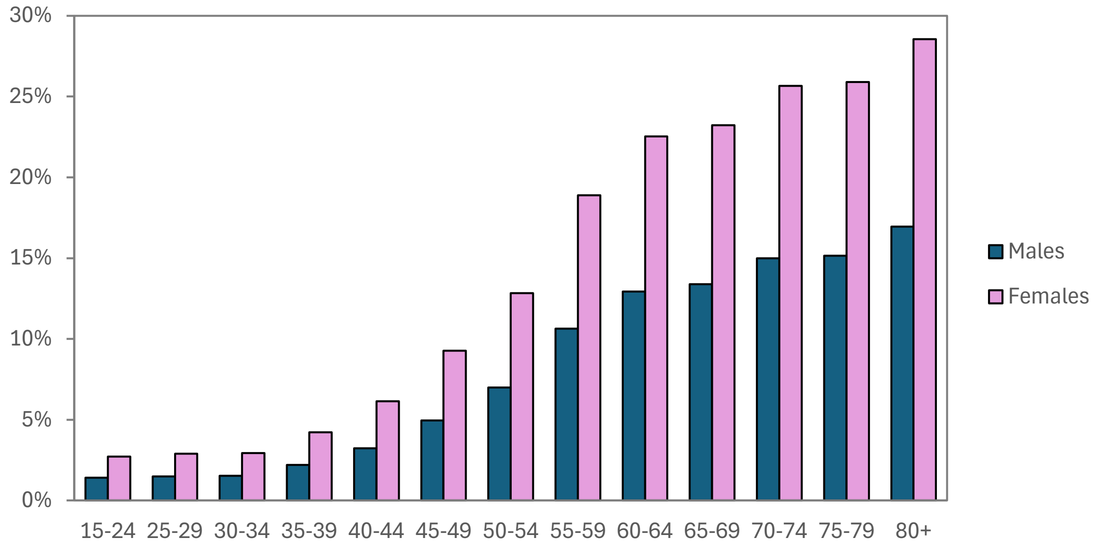

Figure S3: Assumed prevalence of chronic conditions (asthma, arthritis, IHD or stroke)
Prevalence estimates are calculated from a logistic regression model fitted to data from the 2013 General Household Survey data (author's calculations), assuming the effect of sex is constant with respect to age (on the logit scale). IHD = ischaemic heart disease.

The value of $\theta_C$ is difficult to determine precisely. In our meta-analysis of South African studies, we found that the odds ratios relating depression to asthma, arthritis, IHD and stroke were 3.48, 3.17, 3.20 and 4.06 respectively (average of 3.5) [43]. However, we also noted that much of this association could be attributed to confounding by age and other factors. When the analysis was limited to a subset of the data in which we could control for age, the odds ratios for the associations between depression and these four conditions reduced by between 0.3 and

1.2 (average reduction of 0.85) [43]. Based on this, we set $\theta_C$ to 2.5, more conservative than the average unadjusted ratio of 3.5.

##### 1.1.7 The COVID-19 pandemic

International literature, mainly from high-income countries, suggests that the prevalence of depression symptoms increased substantially during 2020, as a consequence of the COVID-19 pandemic [63], and in some settings there have also been increases in recorded diagnoses of depression [64]. Although there are several possible mechanisms that could explain this increase (e.g. increased unemployment and social isolation due to lockdowns, anxiety over the pandemic, loss of loved ones, or the direct effect of 'long COVID'), the two indicators that were found to be most significant in an international meta-regression of prevalence studies over the early phase of the COVID-19 pandemic were the level of mobility (a measure of the severity of lockdowns) and the daily SARS-CoV-2 infection rate [63]. (The excess mortality rate was also a highly significant predictor, but because it was collinearly related to the daily infection rate, it was excluded from the final meta-regression model.) Based on the meta-regression model, the authors estimated that the prevalence of depression in South Africa increased by 43% (95% CI: 30-56%) during 2020, as a result of the COVID-19 pandemic. Although there is some South African data on changes in the depression prevalence over the course of the COVID-19 pandemic [65], it is based on the PHQ-2 screening tool, and thus not directly comparable with the national surveys that were conducted before the COVID-19 pandemic.

In our model, we represent the effect of the COVID-19 epidemic on depression incidence by assuming that depression incidence increases in proportion to the monthly excess mortality. (We lack reliable national data on the extent of changes in mobility over the course of the pandemic, and have therefore chosen to rely only on excess deaths as a measure of the severity of the COVID-19 epidemic.) For each month between January 2020 and December 2022, we assume that if $\lambda_{g,i}(x)$ is the annual incidence of depression in people in risk group $i$, of age $x$ and sex $g$, who have none of the previously mentioned risk factors, then the monthly incidence of depression in all people of risk group $i$, age $x$ and sex $g$ in month $t$ is

$$\lambda_{g,r}(x)(1 + E(t)\theta_D)$$

where $E(t)$ is the number of excess deaths in month $t$ (per 100 000 population) and $\theta_D$ is the proportional increase in depression incidence per unit increase in the monthly excess deaths (per 100 000 population). The excess death rates are those estimated by Bradshaw *et al* [66, 67] from vital registration data, which average 10.9 per month per 100 000 in 2020. If the incidence of depression in South Africa increased by an average of 42.7% in 2020, as estimated by the previous meta-regression model [63], this would suggest a $\theta_D$ value of 0.039 (0.427 / 10.9). This could be an under-estimate, because incidence changes more rapidly than prevalence (and the 42.7% is actually an increase in prevalence), but it could also be an over-estimate, given the limited South African data showing no clear association between the severity of the COVID-19 pandemic and depression symptoms [65]. We therefore represent the uncertainty in the $\theta_D$ parameter using a gamma prior distribution with a mean of 0.039 and a standard deviation of 0.01. This distribution has 2.5 and 97.5 percentiles of 0.022 and 0.061 respectively, and thus allows for a reasonably wide range of uncertainty around the possible magnitude of the effect of the COVID-19 pandemic.

#### 1.2 Duration of depression in the absence of treatment

##### 1.2.1 Heterogeneities in duration

Estimates of the median duration of depression are sensitive to the definition of 'recovery', and many experts distinguish 'remission' (short-term resolution of symptoms) from 'recovery' (a longer-term recovery from symptoms) [68]. For example, Furukawa *et al* [69] found in Japan that the median duration of depression episodes was 2 months if recovery was defined as no depression symptoms for 2 months, whereas the median increased to 4 months if recovery was instead defined as no depression symptoms for at least 6 months. Furukawa *et al* [69] note that the definition of no (or minimal) symptoms for 2 months is the most widely used definition of depression, and we follow this convention. Depression episodes last for a median of 3 months, although this is highly variable [3, 70].

Translating the median duration of 3 months into a mean is complicated by the fact that the distribution of depression durations is highly skewed [71]. If one were to assume the duration of depression symptoms is exponentially distributed, the median of 3 months would correspond to a mean of 4.3 months (3/log(2)). However, the distribution of symptom durations in a Netherlands study [70] was much more highly skewed than an exponential distribution would suggest: although the median duration was 3 months, the Weibull distribution that provides the best fit to the data has a shape parameter of 0.45, which implies a mean duration of 16.8 months. An analysis of Japanese data also found that the exponential model provided a poor fit to the data; the best fit was obtained using a lognormal distribution with a median of 3.2 months and a mean of 6.0 months [71]. We represent the uncertainty in the mean duration of symptoms using a gamma distribution with a mean of 9 months and a standard deviation of 3 months; this distribution has 2.5 and 97.5 percentiles of 4.1 and 15.8 months respectively. The distribution was thus chosen so that its 95% range would roughly cover the range of mean estimates derived from published studies. It is worth noting that our model assumption relates to the duration of untreated disease, but the cited studies measured durations in cohorts who had access to treatment. This could lead to a bias towards under-estimation of the untreated symptom duration. However, it is also possible that selection biases lead to episodes of very short duration being excluded (because clinic-based studies only enrol patients who have sought care, and community-based studies may miss episodes of short duration due to recall bias).

##### 1.2.2 Effects of exercise

In observational studies, physical activity is strongly associated with both lower incidence of depression and higher rates of recovery [12]. South African studies have also found a lower prevalence of depression among people who report regular exercise [14]. Meta-analyses of RCTs of exercise interventions have also found them to be highly effective in reducing symptoms of depression among people with depression [72, 73]. However, we could not identify RCTs that evaluated the effect of exercise on the *incidence* of depression, and therefore chose to model only an effect of exercise on the duration of depression. In this study, we model levels of physical activity in terms of metabolic equivalents of time (MET)-minutes per week. We assume that the duration of depression is proportional to

$$(1-\alpha)^{\log(\tau_g(x))}$$

where $\alpha$ is the proportional reduction in depression symptoms per log increase in $\tau_g(x)$, the number of MET-minutes per week in individuals of sex $g$ and age $x$. We estimate $\alpha$ by simulating the relationship that we might expect to observe between the intensity of exercise interventions (in MET-minutes per week) and the d statistic representing the intervention effect, and comparing this to the results of a meta-regression that assessed the same relationship [72]. We obtain approximate consistency when $\alpha$ is 0.135 (Figure S4), with the model producing d statistics in the range of -0.8 to -0.5, consistent with the results of the meta-regression. We used the log-transformed number of MET-minutes in the above equation in order to ensure that the model matches the relatively weak relationship between the dose of the intervention and its effect.

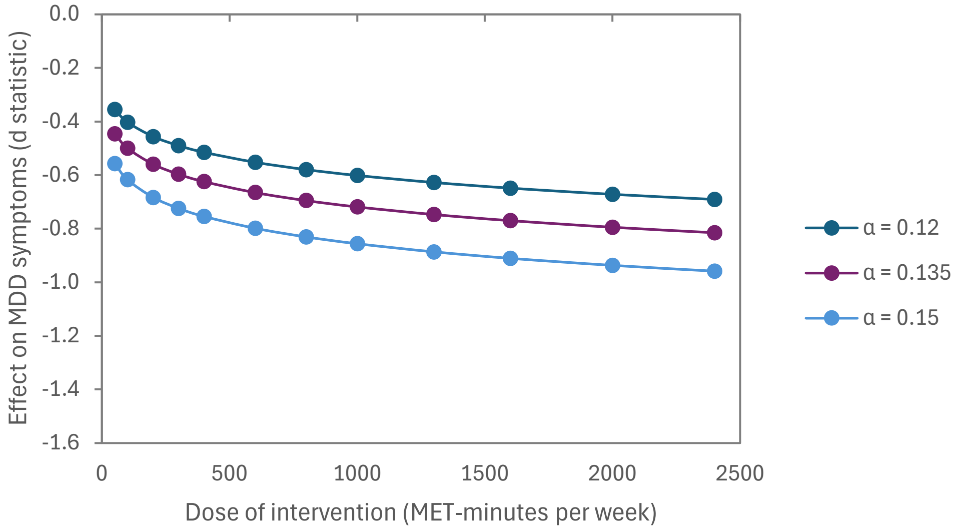

Figure S4: Modelled relationship between intervention intensity and effect of depression symptoms, for different values of $\alpha$

These results are based on simulations of RCTs, each dot representing an RCT comparing depression scores in 1000 individuals before and after an exercise intervention. Baseline MET-minute/week values are sampled from a gamma distribution with mean 1740 and standard deviation 3260 (roughly approximating the distribution in the 2012 SANHANES survey [74]) and baseline numbers of depression symptoms are sampled from a gamma distribution with mean 8.0 and standard deviation 4.8 (based on CESD-10 data from the 2008 NIDS). The d statistic is calculated as the difference between the mean number of depression symptoms before and after the intervention, divided by the standard deviation of the number of depression symptoms at baseline. This is roughly comparable with the Hedges g statistic reported in the meta-regression [72].

We assume levels of physical activity in the South African population consistent with those assumed in the South African Comparative Risk Assessment Study [74]. This study divides the South African population into four physical activity strata (inactive, low active, moderately active and highly active) based on MET-minutes per week. The proportions in the four groups vary by age and sex, and also are assumed to increase between 2003 and 2012. As there have been very few nationally-representative survey estimates of physical activity in South Africa, we adopt a simple approach to model time trends in physical activity. The proportions in each state in the years before 2003 are assumed to be the same as in 2003, the proportions in each state in the years after 2012 are assumed to be the same as in 2012, and for the years between 2003 and 2012 we linearly interpolate between the proportions estimated in 2003 and 2012. The proportions assumed in 2012 are shown in Figure S5.

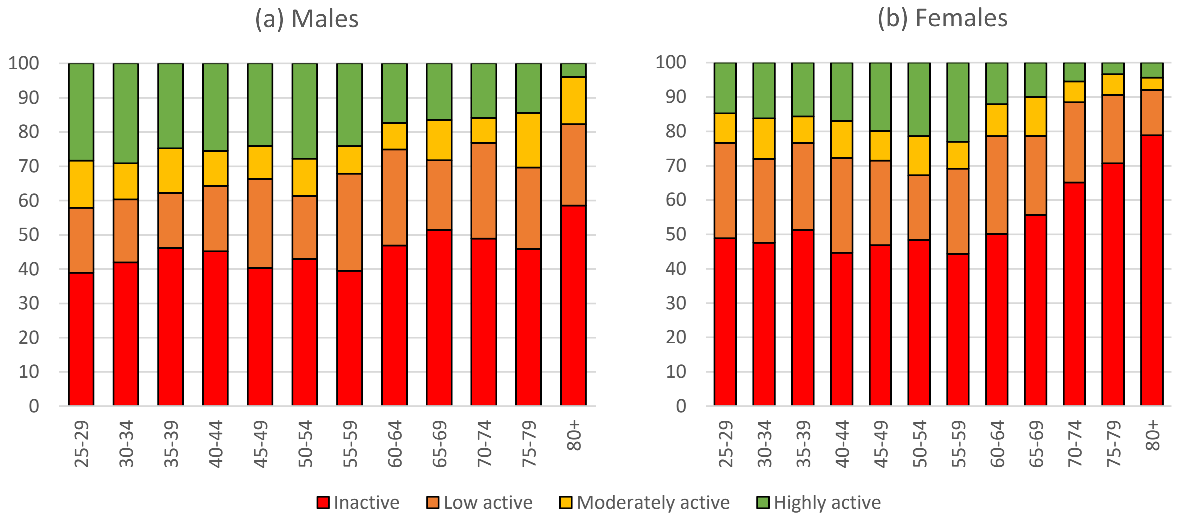

Figure S5: Physical activity levels in 2012
Source: Neethling *et al* [74]

For the purpose of applying the formula for the relative depression duration, we assign median MET-minute/week values to each of the four categories: 50 for inactive, 1500 for 'low active', 6000 for 'moderately active' and 11000 for 'highly active'. (It is worth noting that the median values for the lowest categories are towards the lower end of the interval because the distribution of MET-minutes per week is highly right-skewed.)

#### 1.3 Antidepressant use and effectiveness

Our definition of treatment includes only antidepressant treatment at therapeutic dosage (i.e. excluding people who may be receiving lower dosages for other indications). Sections 1.3.1-1.3.5 describe the assumptions regarding rates of antidepressant initiation, sections 1.3.6-1.3.7 describe the assumptions about the duration of antidepressant use, and section 1.3.8 describes the assumptions about antidepressant effectiveness.

##### 1.3.1 Rate at which women seek care for depression symptoms

In a national survey conducted between 2002 and 2004, 9% of women who had depression in the last 12 months reported having sought mental health care, and this proportion increased to 18.5% if including other healthcare providers (not specifically those providing mental health services) [75]. If it is assumed that depression symptoms in the last 12 months would have lasted for an average of 6 months, then a 9% proportion having sought care implies

$$0.09 = \frac{\gamma}{\gamma + \frac{1}{0.5}}$$

where $\gamma$ is the annual rate of health seeking, and 0.5 is the average duration of depression symptoms. Solving for $\gamma$, we get $\gamma = 0.20$. This could be an under-estimate if we consider that women might also have received mental health care from non-specialist services (if we used

18.5% in place of 9%, we would get $\rho = 0.45$). On the other hand, it could be an over-estimate, since the parameter actually applies to women seeking care for the *first* time, and a higher rate is assumed to apply if women have previously sought mental health care. To represent the uncertainty in the $\gamma$ parameter, we assign a gamma prior with a mean of 0.20 and a standard deviation of 0.1. This distribution has a 97.5 percentile of 0.44, close to the upper bound of 0.45 estimated.

##### 1.3.2 Relative rate of care seeking in men

South African studies suggest that men have lower rates of care seeking for depression symptoms than women. In the nationally representative SASH survey, 9.0% of women who had depression in the last 12 months reported seeking mental health care, compared to only 1.2% of men with depression [75], implying a relative rate of health seeking in men of 0.13. In a study of patients receiving chronic care in the Western Cape, who had symptoms of depression, the relative rate of antidepressant use in men to that in women was 0.31 (95% CI: 0.19-0.49) [76]. In a study of ART patients, however, a relatively high rate of antidepressant use in men was measured relative to that in women, 0.67 (95% CI: 0.65-0.69) [77], though this might be a reflection of the select nature of the sample (men with HIV being less likely to initiate ART than women [78]). Based on these studies, we represent the uncertainty around the relative rate parameter with a gamma distribution, with a mean of 0.30 and standard deviation of 0.15. The mean corresponds roughly to the median of the three studies, while the standard deviation has been chosen such that the 97.5 percentile of the prior distribution (0.66) is close to the estimate in the ART study.

##### 1.3.3 Proportion of people seeking care who are prescribed antidepressants pre-2008

We lack reliable data on the proportion of people seeking care for depression who are prescribed antidepressants. However, access to antidepressants is generally considered to be extremely limited in the public health sector, compared to the private sector, in which antidepressants are more readily prescribed. Since the level of antidepressant use in the public sector was roughly 0.1 times that in the private sector over the 2012-17 period [77], and roughly 30% of South African adults rely on the private health sector for outpatient care [79], it is unlikely that the proportion of people seeking treatment who actually get antidepressants would be more than 0.37 ($0.3 + (1 - 0.3) \times 0.1$, i.e. when optimistically assuming that all people seeking care in the private sector get antidepressants). We therefore represent the uncertainty in this proportion prescribed antidepressants using a beta distribution with a mean of 0.15 and a standard deviation of 0.09. This distribution has a 97.5 percentile of 0.36, close to the upper limit of 0.37.

##### 1.3.4 Relative rate of antidepressant prescription in 2018 and later years (relative to pre-2008)

International studies suggest that the proportion of the population using antidepressants has been increasing over time [80-82]. South African data appear to be consistent with this. For example, Ruffieux *et al* [77] found that among South Africans on ART, the proportion receiving pharmacological treatment for a mental disorder doubled between 2012 and 2017 in private sector patients but increased more modestly in patients using public sector facilities. Oosthuizen *et al* [83] found that between 2009 and 2013, the numbers of people treated with selective serotonin reuptake inhibitors (SSRIs, the most common first-line treatment for depression) increased from 290 000 to 390 000 in the private sector – a 34% increase (although some of this increase may have been due to increasing popularity of SSRIs relative to other antidepressants). In an analysis of antidepressant drug sales data, Alabaku *et al* [82] reported a more modest 16% increase over the 2014-19 period, in South Africa.

It is not clear if these increases represent increasing initiation of antidepressants or increasing duration of antidepressant use. Data from high-income settings suggest much of the increase is driven by increasingly long durations of antidepressant use [84], but corresponding local data are lacking. In the interests of simplicity, we assume the change is driven only by increasing rates of antidepressant initiation (not increasing duration). Since almost all of the available South African data on prevalence of antidepressant use relate to the period 2008-2018, we adopt a parsimonious modelling approach, assuming a linear increase in the rate of antidepressant initiation between 2008 and 2018, but a constant rate of antidepressant initiation in the period before 2008 and a constant (higher) rate in the period after 2018. We represent the uncertainty in the ratio of the post-2018 initiation rate to that pre-2008 using a gamma prior distribution with a mean of 2 and a standard deviation of 0.6. The mean of 2 corresponds roughly to the average increase in the Oosthuizen and Ruffieux public sector data (extended over 10 years). The 2.5 and 97.5 percentiles of the distribution are 1.0 and 3.3 respectively; the former corresponds to a conservative assumption of no change in antidepressant use over time, while the latter is more consistent with the high rate of growth seen in the Ruffieux private sector data.

##### 1.3.5 Relative rate of health seeking if previously treated

Because of heterogeneities in access to healthcare and personal willingness to seek mental healthcare, we assume that individuals who have previously received mental healthcare are more likely to seek mental healthcare than individuals with the same depression symptoms who have not previously been treated. Similar heterogeneities are likely to exist for other diseases; for example, we previously estimated that South African adults who have previously tested HIV-negative are twice as likely to get tested in a year as people who have never previously tested, even after controlling for differences in age, sex and risk behaviour [85]. More extreme differences might be expected in the context of depression care seeking, given that access to mental health care is much more limited than access to HIV care. For example, access to mental health care in users of public primary care is only about 5% of that in private care [77]; if all affected users of private care had previously received treatment and no users of public care had previously received treatment, one would expect the ratio of mental healthcare seeking in the previously-treated to those who were never treated to be 20 (1/0.05). (This is an upper bound because in reality not all affected private sector users have previously been treated, and not all public sector users have never been treated.) We represent the uncertainty in this ratio using a gamma distribution with a mean of 9 and a standard deviation of 5. These parameters were chosen such that the 2.5 and 97.5 percentiles of the distribution (2.0 and 21.1 respectively) would roughly match the noted lower and upper bounds.

##### 1.3.6 Duration of antidepressant use if recently initiated antidepressant treatment

Studies of patients who have recently initiated antidepressant treatment typically find a high rate of treatment discontinuation during the first few months after starting therapy. Studies from high-income countries have found highly variable rates of antidepressant continuation, with the proportion remaining on antidepressant medication 3 months after starting estimated at 28% [86], 54% [87] and 80% [88, 89]. South African private sector data suggest 3-month continuation rates towards the upper end of this range [90], although private sector data may be biased, given that data from other countries suggest better continuation rates in medically insured patients [86, 88]. The range of 28-80% corresponds to an antidepressant duration range of 2.4-13.4 months (e.g. 2.4 = -3/log(0.28)). These estimates could be under-estimates, because monthly discontinuation rates are distorted by the high rate of discontinuation in the first month of treatment [87]. However, they could be over-estimates of the durations that apply while people remain asymptomatic, as many individuals recover while on antidepressants (and we assume there would be longer durations of antidepressant use after recovery). We represent the uncertainty in this parameter using a gamma distribution with a mean of 7 months and a standard deviation of 3 months. This distribution was selected such that the 2.5 and 97.5 percentiles (2.4 and 14.0 months respectively) correspond roughly to the lower and upper limits of the range estimated from international studies.

##### 1.3.7 Duration of antidepressant use if recovered from depression

Very low rates of antidepressant discontinuation are common in people who have been on antidepressants for more than 6 months [91]. In a study of patients receiving chronic care in public health facilities in the Western Cape [76], 82% of those who were receiving a therapeutic dose of antidepressants at baseline were still on antidepressants 14 months later (Naomi Folb, personal communication). This would suggest an average duration of 5.9 years (-1/(log(0.82) × 12/14)). This could be an over-estimate of the average duration, since it does not take into account that some of those who 'remained' on antidepressants might in fact have cycled on and off treatment. However, it could be an under-estimate, as those on antidepressants at baseline included some who were currently experiencing clinical depression (and we assume those individuals would have a shorter duration of antidepressant use). To represent the uncertainty in this parameter, we assign a gamma distribution with a mean of 6 years and a standard deviation of 2 years.

##### 1.3.8 Antidepressant effectiveness

Antidepressants are used in the treatment of both acute and chronic depression. In a systematic review of RCTs, all antidepressant medications were found to increase resolution of acute depression symptoms (with odds ratios varying between 1.4 and 2.1) [92]. Based on this systematic review, we assume that in people receiving antidepressants the rate of recovery from depression is 1.8 times that in people with untreated depression.

An early meta-analysis suggested that antidepressant treatment could reduce the rate of relapse by around 70% [93]. However, more recent meta-analyses suggest that antidepressant treatment is similarly effective when compared against cognitive behavioural therapy (CBT) in preventing relapse [94, 95], which suggests a more moderate effect (systematic reviews estimate a roughly 30% reduction in incidence due to CBT [94, 96]). Based on the latter, we assume that individuals who are currently using antidepressants have a 30% lower rate of recurrence than people who are not currently using antidepressants.

#### 1.4 Calibration to depression prevalence data

We adopt a Bayesian approach to model calibration and uncertainty analysis. This involves specifying prior distributions to represent plausible ranges of uncertainty around key model parameters (inputs), likelihood functions that represent the degree of consistency between the model outputs and the calibration targets, and the calculation of posterior distributions that represent the parameter combinations most consistent with both the prior distributions and calibration data. The sections that follow describe each of these steps.

It is worth noting that we follow two separate Bayesian calibration procedures. The first step (described in section 1.4) involves calibration to national depression prevalence survey estimates. The second step (described in section 1.5) involves calibration to data on antidepressant use.

##### 1.4.1 Prior distributions

Table S3 summarizes the parameters that are varied in the calibration to the depression prevalence data, and the prior distributions assigned to each. Most of these prior distributions have been justified previously (and the relevant section of the supplementary materials is referenced in the final column). Some of the prior distributions are described further below (section 1.4.2).

Table S3: Prior distributions in the calibration to depression prevalence survey data

| Parameter | Symbol | Prior | Mean | SD | Ref |
|---|---|---|---|---|---|
| Mean duration of untreated symptoms | $D$ | Gamma (9, 1) | 9 | 3 | 1.2.1 |
| Incidence of depression in low-risk men | $\lambda_0$ | Gamma (7.29, 270) | 0.027 | 0.010 | 1.1.1 |
| Incidence of depression in low-risk women | $\lambda_1$ | Gamma (6.84, 201) | 0.034 | 0.013 | 1.1.1 |
| Ratio of incidence in low-risk group to high-risk group | $\Omega$ | Beta (1.28, 2.98) | 0.3 | 0.2 | 1.1.1 |
| Increase in depression incidence per 10-year increase in age (RR) | $B$ | Gamma (100, 100) | 1 | 0.1 | 1.1.2 |
| % increase in depression incidence per unit excess deaths (per 100,000) | $\theta_D$ | Gamma (15.2, 390) | 0.039 | 0.01 | 1.1.7 |
| Ratio of point prevalence to 12-month prevalence, using CIDI | $R_1$ | Beta (20.5, 6.13) | 0.77 | 0.08 | 1.4.2 |
| Ratio of true prevalence to PHQ-9 prevalence | $R_2$ | Gamma (40.5, 116) | 0.35 | 0.055 | 1.4.2 |
| Ratio of true prevalence to CES-D prevalence | $R_3$ | Gamma (40.5, 116) | 0.35 | 0.055 | 1.4.2 |
| Effect of log(prevalence) on association between initial and later depression | $\beta$ | Normal (-0.16, 0.04) | -0.16 | 0.04 | 1.4.3 |
| RR of depression incidence in 2025, relative to 2020* | - | Gamma (25.0, 25.0) | 1 | 0.2 | -* |
| Incidence of depression in high-risk men† | - | Gamma (1, 0.571) | 1.75 | 1.75 | 1.1.1 |
| Incidence of depression in high-risk women† | - | Gamma (1, 0.571) | 1.75 | 1.75 | 1.1.1 |

CES-D = Centre for Epidemiological Studies Depression Scale. CIDI = Composite International Diagnostic Interview, PHQ-9 = Patient Health Questionnaire-9, RR = relative rate, SD = standard deviation. * Only relevant to Model B (described in the main text). † Only relevant to Model D (described in the main text).

#### 1.4.2 Likelihood function: cross-sectional prevalence data

We rely on data from nine nationally-representative surveys that measured the prevalence of depression:

- The 2002-2004 Stress and Health Survey (SASH) [97], which relied on the CIDI diagnostic interview;
- The National Income Dynamics Study (2008, 2010-11, 2012, 2014-15 and 2017) [98], which relied on the CES-D-10 screening tool; and
- The South African Human Development Pulse Surveys conducted in 2021, 2022 and 2024 [17], which relied on the PHQ-9 screening tool.

In all cases, prevalence estimates are calculated using survey weights, and taking into account the relevant survey design.

Although the CIDI is considered a diagnostic interview rather than a screening tool, it is nevertheless based on a fully structured interview, which is not as accurate as the semi-structured SCID, the widely used gold standard in depression diagnosis. In comparisons of the CIDI and SCID, the CIDI on average gives a lower prevalence, although this was not significantly different from 1 in a synthesis of estimates from individual patient data meta-analyses (aOR 0.83, 95% CI: 0.54-1.27) [99]. The CIDI tends to over-diagnose depression when there are few symptoms, but under-diagnoses when symptoms are more frequent [99]. In the SASH survey, the CIDI was used to report a 12-month prevalence of depression, rather than an estimate of the point prevalence of depression. The ratio of the former to the latter, based on an international meta-regression of prevalence surveys, is 1.3 (95% CI: 1.1-1.6) [8]. We specify a beta prior distribution to represent the uncertainty in the inverse of this ratio ($R_1$): the prior distribution has a mean of 0.77 (equal to 1/1.3) and a standard deviation of 0.08. With these parameters the prior distribution has 2.5 and 97.5 percentiles of 0.60 and 0.91 respectively, close to the inverses of the 95% confidence interval (1.1-1.6).

When calibrating to the NIDS data (2008-2017), it is important to take into account that the CES-D-10 has limited specificity. In previous analyses of NIDS data, investigators have commonly used cutoffs of 10+ to define probable depression [15, 18, 100]. At this threshold, the CES-D-10 is estimated to have sensitivity of 81-86% and specificity of 58-87% across different ethnic groups in South Africa [101]. This is roughly consistent with the more complete CES-D screening tool, which is estimated to have an average sensitivity of 83% and average specificity of 78% at a cutoff of 20+ [6] (roughly comparable to the 10+ threshold for CES-D-10 because it uses 20 questions instead of 10). These sensitivity and specificity estimates are based mostly on studies that used fully-structured interviews to diagnose depression (rather than semi-structured interviews, which are generally considered the gold standard), and this may lead to some under-estimation of sensitivity, although specificity appears to be relatively unaffected [102]. Although it may seem natural to specify prior distributions for the sensitivity and specificity parameters, we would expect sensitivity and specificity parameters to be highly variable between sub-populations, whereas the ratio of the true prevalence to the measured prevalence (using CES-D-10) is expected to be more stable (see Appendix B). Based on this, we specify a gamma prior on the ratio of true prevalence to measured prevalence ($R_3$), with a mean of 0.35 and standard deviation of 0.055. The Global Burden of Disease study adopts a similar approach, specifying a 'predictive validity' factor that represents the ratio of true prevalence to the 'probable disorder prevalence' [63]. The mean and standard deviation of the prior distribution were selected based on a systematic review and meta-analysis of depression prevalence studies [8], which estimated that the odds ratio comparing 'true' depression prevalence to that estimated based on symptom screening tools (like CES-D-10) was 0.34 (95% CI: 0.25-0.48). The prior standard deviation of 0.055 was selected such that the 2.5 and 97.5 percentiles of the prior distribution (0.25 and 0.47 respectively) would be approximately equal to the 95% confidence interval limits from the meta-analysis.

Similarly, when calibrating to the 2021-2024 Human Development Pulse Survey data, it is important to take into account that the PHQ-9 has limited specificity. When using a threshold of 10+ to define depression (as in the primary analysis of the 2021 survey data [17]), the average sensitivity of the PHQ-9 is 0.88 and the average specificity is 0.85 [102]. The results from this international meta-analysis are different from those found in a South African validation study, which estimated a substantially lower sensitivity (57-65%) but similar specificity (85-95%) [101]. This lower sensitivity is likely to be because the South African study used a fully structured interview to diagnose depression, and this tends to be associated with poorer sensitivity [102]. As with the CES-D-10, we specify a ratio of the true depression prevalence to that measured by the PHQ-9 ($R_2$), and we represent the uncertainty in this ratio using a gamma prior distribution with a mean of 0.35 and standard deviation of 0.055.

Unpublished South African data suggest that the relative accuracy of the PHQ-9 and CES-D-10 may differ between men and women. Table S4 compares the estimates of the $R_2$ and $R_3$ ratios when calculated for males and females separately. The ratio is consistently lower in males than in females, with the male ratios being consistently around 0.8 times the female ratios. This suggests that symptom screening tools overstate the true prevalence by proportionally more in males compared to females. To take this into account, we multiply the $R_2$ and $R_3$ ratios by sex-specific adjustment factor $\Psi_g$, which is 0.86 for males and 1.08 for females. These values were chosen such that the male-to-female ratio is 0.8 and the average ratio is close to 1 (taking into account the higher prevalence of depression in women).

Table S4: Sex-specific ratios of diagnosed prevalence (based on MINI) to prevalence based on symptom screening

| Study | Screening tool | Sex | Diagnosed prevalence | Symptom prevalence | Ratio |
|---|---|---|---|---|---|
| Baron *et al* [101] | CES-D-10 | M | 6.8% | 17.5% | 0.390 |
| | | F | 12.3% | 25.3% | 0.486 |
| | PHQ-9 | M | 6.8% | 19.2% | 0.356 |
| | | F | 12.3% | 27.5% | 0.447 |
| Tlali *et al* [59] | PHQ-9 | M | 7.2% | 19.9% | 0.364 |
| | | F | 15.8% | 35.5% | 0.446 |

Data are unpublished (personal communications with Emily Baron and Andreas Haas).

Having specified the prior distributions for the 'bias' parameters, we now define the mathematical form of the likelihood. We define $M_g(x,t)$ as the model estimate of depression prevalence in year $t$, in people of sex $g$ and age $x$. Similarly, $S_{g,i}(x,t)$ is the survey estimate of depression prevalence in year $t$, in people of sex $g$ and age $x$, based on screening/diagnostic tool $i$. Then we calculate the likelihood for $S_{g,i}(x,t)$ by assuming that

$$\log\left(\frac{S_{g,i}(x,t)}{1-S_{g,i}(x,t)}\right) = \log\left(\frac{M_g(x,t)/R_i}{1-M_g(x,t)/R_i}\right) + \varepsilon_g(x,t)$$

where $\varepsilon_g(x,t)$ is the random error term (representing sampling variability in measured depression prevalence) and we assume $\varepsilon_g(x,t) \sim N\left(0, \sigma_g^2(x,t)\right)$. The variance of the random error term, $\sigma_g^2(x,t)$, is calculated from the 95% confidence intervals around each survey prevalence estimate (after logit transformation of the limits).

In the case of the prevalence data from the NIDS and Human Development Pulse Surveys, we adjust the above formula to take into account the assumed sex differences in the accuracy of the screening tools:

$$\log\left(\frac{S_{g,i}(x,t)}{1-S_{g,i}(x,t)}\right) = \log\left(\frac{M_g(x,t)/\left(R_i\Psi_g\right)}{1-M_g(x,t)/\left(R_i\Psi_g\right)}\right) + \varepsilon_g(x,t)$$

##### 1.4.3 Likelihood function: associations between initial and later depression prevalence (longitudinal data)

Although it is important to quantify depression prevalence (as described in the previous section), it is also important to assess heterogeneity in depression incidence. This is because the cost-effectiveness and relative impact of different interventions is likely to depend on this heterogeneity. For example, if there is a small subset of the population that accounts for a high proportion of all depression episodes (e.g., due to genetic factors), it may be most cost-effective to target counselling and therapeutic interventions to this sub-population. On the other hand, if there is less heterogeneity attributable to innate factors, and variation in depression incidence is more attributable to external factors such as violence exposure and alcohol, it may be relatively more cost-effective to focus on interventions that address these risk factors.

Unfortunately it is difficult to quantify heterogeneity in depression incidence. However, we can approximate it by assessing the association between initial depression and later depression, in the context of longitudinal studies – assuming measurements are taken sufficiently far apart, and assuming that the average duration of symptoms is short relative to this measurement delay. We define this association in terms of an odds ratio: if the odds ratio is close to 1, it implies that there is relatively little heterogeneity in depression incidence, whereas an odds ratio substantially greater than one implies heterogeneity in depression incidence.

We calculate these odds ratios using data from the NIDS surveys mentioned in the previous section. There are five waves, and we consider the odds ratios from four inter-wave comparisons, calculated from consecutive waves (each roughly two years apart) in the subset of adults who had depression symptoms measured across both waves. Results are shown in Figure S6, using four possible thresholds for defining depression (although a CES-D-10 threshold of 10 or higher is usually considered defining of 'probable depression', the CES-D-10 screening tool has low specificity, and thus using a higher threshold might more accurately reflect the 'true' prevalence of depression). The results suggest that both in males and females, higher thresholds are associated with higher odds ratios – though there is also a corresponding loss of precision at higher thresholds.

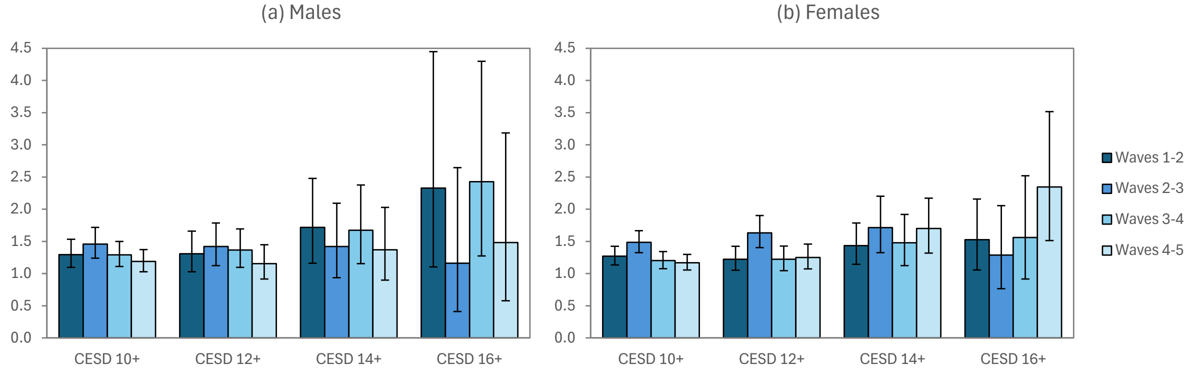

Figure S6: Odds ratios for the association between depression at the start and end of each inter-survey period

Higher CES-D-10 thresholds correspond to lower levels of depression prevalence. We therefore fit meta-regression models to these data, to assess the extent to which the variation in odds ratios is explained by differences in the prevalence of depression in the first survey. The log of the depression prevalence is significantly negatively associated with the log of the odds ratio ($\beta$ = -0.155, 95% CI: -0.240 to -0.069) in a simple univariable model, and similar results are obtained when controlling for sex, period and the sex-period interaction ($\beta$ = -0.165, 95% CI: -0.244 to -0.087). Based on this, we calculate the likelihood by comparing the modelled odds and survey odds after adjusting for differences in prevalence:

$$\log\left(O_g^s(t)\right) = \log\left(O_g^m(t)\right) + \beta\left(\log\left(W_g^s(t)\right) - \log\left(W_g^m(t)\right)\right)$$

where $O_g^s(t)$ is the inter-survey odds ratio for the association between depression at time $t$ and that at the next survey (approximately time $t + 2$), in adults of sex $g$, $O_g^m(t)$ is the corresponding model odds ratio, $W_g^s(t)$ is the survey prevalence of depression in adults of sex $g$ at time $t$, and $W_g^m(t)$ is the corresponding model estimate of depression. We calculate the likelihood assuming that the difference between the log odds ratio in the survey (the left-hand side of the equation) and the adjusted log odds ratio from the model (the right-hand side of the equation) is normally distributed with zero mean and standard deviation calculated from the survey log odds ratio.

Two points require further explanation. Firstly, the $\beta$ term is the same as that estimated in the meta-regression. We represent the uncertainty in this parameter using a normal prior distribution with a mean of -0.16 and a standard deviation of 0.04. Secondly, it would be incorrect to calculate the likelihood for all possible CES-D thresholds, as this would imply double-counting the same survey data. To avoid this, we use only the CES-D threshold of 14 when calculating $O_g^s(t)$ and $W_g^s(t)$. Although there is no 'right' threshold, we chose this as a compromise between a low threshold (which will be associated with an inflated prevalence and a correspondingly large adjustment) and a high threshold (which is associated with imprecise survey odds ratios, as shown by the wide confidence intervals for the 16+ threshold in Figure S6).

We calculate the model odds ratio $O_g^m(t)$ as

$$O_g^m(t) = \frac{N_{g,0,0}(t)N_{g,1,1}(t)}{N_{g,0,1}(t)N_{g,1,0}(t)}$$

where $N_{g,d,j}(t)$ is the number of people of sex $g$ and depression status $d$ at time $t$, who are expected to have depression status $j$ two years later (depression status is 0 if not depressed and 1 if depressed). This is the standard contingency table approach to calculating the odds ratio [103]. The terms in this equation are in turn calculated as

$$N_{g,d,1}(t) = \sum_x \sum_h \sum_i N_{g,d,h,i}(x,t)Y_{g,d,h,i}(x,t)$$
$$N_{g,d,0}(t) = \sum_x \sum_h \sum_i N_{g,d,h,i}(x,t)\left(1 - Y_{g,d,h,i}(x,t)\right)$$

where $N_{g,d,h,i}(x,t)$ is the number of people of sex $g$, age $x$ and depression status $d$ at time $t$, who are in risk group $i$ and in HIV state $h$; and $Y_{g,d,h,i}(x,t)$ is the proportion of this group who are expected to be depressed in two years' time. If someone is depressed at time $t$, the probability that they will be depressed $z$ years later is

$$Y_{g,1,h,i}(x,t) = 1 - \frac{\rho_{g,h,i}(x,t)\left(1 - \exp\left(-\left(\lambda_{g,h,i}(x,t) + \rho_{g,h,i}(x,t)\right)z\right)\right)}{\lambda_{g,h,i}(x,t) + \rho_{g,h,i}(x,t)}$$

where $\lambda_{g,h,i}(x,t)$ and $\rho_{g,h,i}(x,t)$ are the annual depression incidence and recovery rates respectively. Similarly, if someone is *not* depressed at time $t$, the probability that they will be depressed $z$ years later is

$$Y_{g,0,h,i}(x,t) = \frac{\lambda_{g,h,i}(x,t)\left(1 - \exp\left(-\left(\lambda_{g,h,i}(x,t) + \rho_{g,h,i}(x,t)\right)z\right)\right)}{\lambda_{g,h,i}(x,t) + \rho_{g,h,i}(x,t)}$$

These expected prevalence levels are thus calculated from the modelled rates of depression incidence and recovery in each age/sex/risk/HIV category.

##### 1.4.4 Posterior distributions

We use Incremental Mixture Importance Sampling [104] to numerically approximate the posterior distribution. This is an iterative sampling procedure, with each sampling step sampling more intensively from the subset of the parameter space in which the posterior distribution is maximized. We follow these steps:

1. In the first IMIS step, we randomly sample 10 000 parameter combinations from the prior distributions described in section 1.4.1.
2. For each parameter combination, we run the model and use the model outputs to calculate a likelihood statistic (the product of the likelihoods defined in sections 1.4.2 and 1.4.3).
3. A new sampling distribution is defined based on the subset of parameter combinations that have the highest likelihood values, and another sample is drawn from the new sampling distribution (sample size 1000), with likelihood values being similarly calculated for each parameter combination.
4. We calculate the expected fraction of posterior parameter combinations that would be unique if we were to draw a weighted sample of 1000 parameter combinations from the combinations generated thus far (using appropriate posterior estimates and sampling probabilities as weights).
5. We repeat steps 3 and 4 until the expected fraction unique is greater than $1 - e^{-1}$ (0.63). We then draw a random sample of 1000 parameter combinations, using the same weights as described in the previous step, and this constitutes the posterior sample.

#### 1.5 Calibration to antidepressant use data

For the purpose of calibrating the model to antidepressant data, we keep the depression incidence and recovery parameters fixed at the posterior means calculated in the previous calibration step (described in section 1.4).

##### 1.5.1 Prior distributions

Table S5 summarizes the prior distributions used to represent the uncertainty in the antidepressant parameters. Most of these parameters and prior distributions have been described previously (referenced in the final column of the table). However, three of the parameters are used only in the model calibration process, and are described further below.

Table S5: Prior distributions in the calibration to depression prevalence survey data

| Parameter | Symbol | Prior | Mean | SD | Ref |
|---|---|---|---|---|---|
| Annual health seeking rate in women | $\gamma$ | Gamma (4, 20) | 0.2 | 0.1 | 1.3.1 |
| RR of health seeking in men | $K$ | Gamma (4, 13.3) | 0.3 | 0.15 | 1.3.2 |
| Proportion prescribed antidepressants pre-2008 | $\eta$ | Beta (2.21, 12.5) | 0.15 | 0.09 | 1.3.3 |
| RR of antidepressants in 2018 | $J(.)$ | Gamma (11.1, 5.56) | 2 | 0.6 | 1.3.4 |
| RR of health seeking if previously treated | $P$ | Gamma (3.24, 0.36) | 9 | 5 | 1.3.5 |
| Average months on antidepressants while experiencing symptoms | $12/\delta_0$ | Gamma (5.44, 0.78) | 7 | 3 | 1.3.6 |
| Average years on antidepressants post-recovery | $1/\delta_1$ | Gamma (9, 1.5) | 6 | 2 | 1.3.7 |
| RR of antidepressant use in public sector | $U$ | Beta (8.10, 81.9) | 0.09 | 0.03 | 1.5.1 |
| RR of antidepressants in people receiving treatment for other chronic conditions | $\chi$ | Gamma (9, 3) | 3.00 | 1.00 | 1.5.1 |
| Coefficient of variation in random effects | $\mu$ | Uniform (0, 1) | 1.00 | 0.29 | 1.5.1 |

RR = relative rate, SD = standard deviation.

*Relative rate of antidepressant prescription in the public sector*

In a study of patients receiving ART in South Africa, Ruffieux *et al* [77] estimated the relative rate of antidepressant use among those using public primary care facilities was only 0.06 times that in the private sector. A higher relative rate of antidepressant use was measured in those using public tertiary care (aRR 0.48, again relative to private care), but this is less likely to be representative, as only 6% of public sector users were receiving care from tertiary facilities. Averaged across primary and tertiary care users, the relative rate in the public sector is around 0.09 times that in the private sector. We represent the uncertainty in this parameter using a beta prior distribution with a mean of 0.09 and a standard deviation of 0.03.

*Relative rate of antidepressant prescription in people treated for chronic conditions*

Many studies on the prevalence of antidepressant use are conducted in patients who are currently treated for other long-term/chronic conditions. These may over-estimate the true prevalence of antidepressant use in the general population, (a) because people who attending healthcare services regularly for other conditions may find it easier to access antidepressant medication, and (b) because several chronic conditions are associated with an increased risk of depression [43]. Most studies that have evaluated the association between antidepressant use and treatment of other conditions have produced varying results. Two studies in the Netherlands found odds ratios for the association between antidepressant use and treatment of other chronic conditions of 1.38 [105] and 1.83 [106]. In a Brazilian study, receipt of antidepressants was significantly associated with receiving 2-4 other medications (aOR 4.17, 95% CI: 3.16-5.51) or more than 4 other medications (aOR 6.30, 95% CI: 4.42-9.00) [107]. (This is likely to be an over-estimate of the odds associated with receiving only one other medication.) Although we lack comparable South African evidence, one South African study found that people with hypertension were significantly more likely to be on treatment if they were receiving ART (aOR 1.63, 95% CI: 1.21-2.19), and a similar association was seen in diabetes patients (although this was not significant) [108]. This suggests that people in HIV care are more likely to receive screening and treatment for other conditions, and the same is likely to be true for the effect of receiving treatment for other chronic conditions on access to antidepressant treatment. To represent the uncertainty in the relative rate of antidepressant rate in people with other chronic conditions, we assign a gamma prior distribution with a mean of 3 and standard deviation of 1. This distribution was chosen such that the 2.5 percentile (1.37) would correspond to the value estimated from the first Netherlands study and the 97.5 percentile (5.25) would correspond roughly to the upper limit estimated based on the Brazilian data.

*Coefficient of variation for random effects*

In calculating the likelihood for the antidepressant prevalence data, we include a random effect term, to account for sources of heterogeneity not included in our model (e.g. due to differences across locations, methods used to elicit antidepressant use, or sampling methods). In calibrating the model we specify a coefficient of variation parameter, which represents the ratio of the standard deviation of the random effects to the 'true' prevalence of antidepressant use (as predicted by the model). One crude approach to estimating this coefficient of variation is to estimate it directly from the data used in calibration: across the eight populations for which we have antidepressant prevalence estimates, the average prevalence is 6.4% and the standard deviation is 6.7%, which implies a coefficient of variation of 1.05. However, this is almost certainly an upper bound on the true coefficient of variation, as it ignores the possibility that some of the variation may be explained by factors already implicit in our model (e.g. public-private differences and changes over time) and simple binomial variation. We represent the uncertainty in the coefficient of variation using a uniform (0, 1) distribution. This distribution was chosen to reflect the assumption that the true value is unlikely to be above 1, but there is substantial uncertainty regarding the true value; we therefore consider all values between 0 and 1 to be equally probably, *a priori*.

##### 1.5.2 Likelihood function: survey and clinical cohort data

There are two types of antidepressant use data that we use in calibrating our model: surveys or clinical cohort studies that collect individual-level data on proportions of people using antidepressants, and drug distribution data, which provide information on the volume of antidepressants dispensed in a population, over a certain period. Studies were identified by performing PubMed and Google searches using the terms 'antidepressant' and 'South Africa', and by consulting local collaborators. The method for calculating the likelihood is specified separately for each data source; this section describes the approach for the survey data, and the next section (1.5.3) describes the approach for the drug distribution data.

Table S6 summarizes the studies identified. One study was excluded [109] because the same data were reported more fully in another publication [77]. Another study was excluded because it reported the cumulative number of people receiving antidepressants over an extended period, and it was not possible to convert this into a 'point prevalence' of antidepressant use [110].

Table S6: South African studies on the prevalence of antidepressant use

| Study | Year | % private | % male | % treated for chronic | n | % on AD |
|---|---|---|---|---|---|---|
| Folb *et al* [76] | 2011 | 0% | 27.3% | 100% | 4363 | 9.0% |
| Spies *et al* [5] | 2011 | 0% | 0% | 67% | 148 | 0.0% |
| Zani *et al* [111] | 2015 | 0% | 18.2% | 100% | 2002 | 0.1% |
| Petersen *et al* [112] | 2015 | 0% | 18.1% | 100% | 1043 | 1.7% |
| Welthagen & Els [113] | 2008 | 100% | 61.9% | 19% | 15664 | 18.3% |
| Peltzer *et al* [114] | 2011 | 0% | 54.5% | 100% | 4900 | 9.3% |
| Ruffieux *et al* [77] | 2012 | 100% | 34% | 100% | 17274 | 7.2% |
| Ruffieux *et al* [77] | 2013 | 100% | 34% | 100% | 36749 | 8.2% |
| Ruffieux *et al* [77] | 2014 | 100% | 34% | 100% | 43110 | 13.1% |
| Ruffieux *et al* [77] | 2015 | 100% | 34% | 100% | 48346 | 13.6% |
| Ruffieux *et al* [77] | 2016 | 100% | 34% | 100% | 51250 | 14.9% |
| Ruffieux *et al* [77] | 2017 | 100% | 34% | 100% | 23715 | 14.9% |
| Ruffieux *et al* [77] | 2012 | 0% | 29% | 100% | 10228 | 0.7% |
| Ruffieux *et al* [77] | 2013 | 0% | 29% | 100% | 19667 | 1.0% |
| Ruffieux *et al* [77] | 2014 | 0% | 29% | 100% | 20364 | 1.0% |
| Ruffieux *et al* [77] | 2015 | 0% | 29% | 100% | 19147 | 1.0% |
| Ruffieux *et al* [77] | 2016 | 0% | 29% | 100% | 16415 | 1.1% |
| Ruffieux *et al* [77] | 2017 | 0% | 29% | 100% | 5463 | 1.1% |

AD = antidepressants.

In total, 18 prevalence data points were identified from 7 studies. Seven of the 18 data points related to the private sector, and all but two data points related to individuals who were receiving treatment for other chronic conditions (mostly HIV). Eight of the data points were from the Western Cape [5, 76, 77], but all of the private sector estimates were from national datasets.

For the purpose of standardizing the comparisons between the model estimates and the survey/cohort estimates of antidepressant use, it was necessary to include covariate information, which was not always reported. To fill these gaps, we made the following assumptions:

- Where the period was reported as an interval, we took the average of the start and end years (rounding down to the lowest integer).
- The proportion using private healthcare was not reported in one study [113], but it was set to 100% since this was a workplace survey that sampled mostly white-collar workers. This study also did not report the proportion treated for chronic conditions, but this was estimated from the 2009 General Household Survey (for adults who are medical scheme members).
- The denominators were not reported for the prevalence estimates of Ruffieux *et al* [77], but were approximated based on the 95% confidence interval widths.

We calculate the likelihood for each data point using a beta-binomial distribution: the binomial component takes account of the sample size, while the beta component reflects the random effects (variation between studies that is not accounted for by binomial variation or other covariates already included in the model).

To provide more detail, suppose that $M(g, t)$ represents the model estimate of the prevalence of antidepressant use in adults of sex $g$ in year $t$. We define a corresponding 'base' rate of antidepressant use in private sector users, who are not receiving treatment for any other chronic conditions, $B(g,t)$ (the choice of 'base' category is arbitrary). We estimate the base rate by noting that

$$M(g,t) = \frac{\sum_h \sum_c w_{g,t}(h,c)B(g,t)U^h\chi^c}{\sum_h \sum_c w_{g,t}(h,c)}$$

where $w_{g,t}(h,c)$ is the size of the adult population (aged 15+) of sex $g$ in year $t$, who use health sector $h$ (0 for private, 1 for public) and have chronic medication status $c$ (1 if receiving medication for other chronic conditions, 0 otherwise); $U$ is the relative rate of antidepressant use in the public health sector; and $\chi$ is the relative rate of antidepressant use in people receiving treatment for other chronic conditions. From the previous equation it follows that

$$B(g,t) = \frac{M(g,t)\sum_h \sum_c w_{g,t}(h,c)}{\sum_h \sum_c w_{g,t}(h,c)U^h\chi^c}$$

The $w_{g,t}(h,c)$ parameters are estimated from the 2009 to 2015 General Household Surveys, which included questions about receipt of treatment for chronic conditions (asthma, diabetes, cancer, HIV, hypertension, arthritis, stroke or heart problems). (For years before 2009, we use the 2009 data as weights, and for years after 2015 we use the 2015 data as weights – since the earlier and later surveys did not include these questions about chronic medication use.) The $U$ and $\chi$ parameters are varied in the model calibration process, and the prior distributions used to represent the uncertainty in these parameters were specified previously.

Having calculated the $B(g,t)$ terms, we then calculate $A_i(t_i)$, the adjusted model estimate of the prevalence of antidepressant use we might *expect* to observe in study $i$ (conducted in year $t_i$), based on the covariate distribution in study $i$. Mathematically, this is

$$A_i(t_i) = \left[p_m(i)B(0,t_i) + \left(1 - p_m(i)\right)B(1,t_i)\right]\left(p_h(i) + \left(1 - p_h(i)\right)U\right) \times \\ \left(p_c(i)\chi + \left(1 - p_c(i)\right)\right)$$

where $p_m(i)$ is the proportion of study $i$ participants who are male, $p_h(i)$ is the proportion who use the private sector, and $p_c(i)$ is the proportion receiving treatment for other chronic conditions. These study-specific proportions are the values specified in Table S6. We make the simplifying assumption that the distributions across sex and chronic medication use are mutually independent, as most studies do not provide enough detail to calculate the joint distribution across sex and chronic medication use.

Now suppose that $G_i$ is the 'true' prevalence in study $i$ (if we were to repeat the same study in the same population infinitely many times, removing binomial variation), i.e. adding to $A_i(t_i)$ the random effect described previously. We assume that $G_i$ is beta-distributed with mean $A_i(t_i)$ and standard deviation $A_i(t_i)\mu$ (i.e. $\mu$ is the coefficient of variation). If $a_i$ and $b_i$ are the parameters of the beta distribution, we estimate these using the method of moments:

$$a_i = A_i(t_i)\left(\frac{1 - A_i(t_i)}{A_i(t_i)\mu^2} - 1\right)$$

$$b_i = \left(1 - A_i(t_i)\right)\left(\frac{1 - A_i(t_i)}{A_i(t_i)\mu^2} - 1\right)$$

Finally, assume that in study $i$, $x_i$ is the number of people found to be using antidepressants out of a total sample of $n_i$ individuals (determined from the last two columns in Table S6). We calculate the binomial likelihood by integrating over all possible values of $G_i$, using the beta density function as weights. This means that the likelihood is

$$L(x_i|a_i, b_i, n_i) = \int_0^1 \binom{n_i}{x_i} G_i^{x_i}(1 - G_i)^{n_i - x_i} \frac{G_i^{a_i}(1 - G_i)^{b_i}}{\mathrm{Beta}(a_i, b_i)} dG_i$$
$$= \frac{\Gamma(n_i + 1)\Gamma(a_i + b_i)\Gamma(x_i + a_i)\Gamma(n_i - x_i + b_i)}{\Gamma(x_i + 1)\Gamma(n_i - x_i + 1)\Gamma(a_i)\Gamma(b_i)\Gamma(n_i + a_i + b_i)}$$

where the final expression is the standard likelihood for a beta-binomial distribution.

A complication arises when there are multiple measurements (for different years/sectors) in the same study, as in the study of Ruffieux *et al.* In this case, it would be incorrect to apply the same formulas because that would imply the random effects from the single study are statistically independent, when in fact they are highly correlated. Our approach is therefore to calculate an average random effect adjustment factor, $q_i$, which is calculated as

$$q_i = \frac{1}{12}\sum_{j=1}^{12} \frac{x_{ij}/n_{ij}}{A_{ij}(t_{ij})}$$

where the '$ij$' subscript represents the $j^{\text{th}}$ estimate from study $i$ (in the case of the Ruffieux *et al* study, there are 12 prevalence estimates). Then the parameters of the beta distribution are calculated as

$$a_{ij} = A_{ij}(t_{ij})q_i\left(\frac{1 - A_{ij}(t_{ij})q_i}{A_{ij}(t_{ij})q_i\tilde{\mu}^2} - 1\right)$$
$$b_{ij} = \left(1 - A_{ij}(t_{ij})q_i\right)\left(\frac{1 - A_{ij}(t_{ij})q_i}{A_{ij}(t_{ij})q_i\tilde{\mu}^2} - 1\right)$$

where $\tilde{\mu}$ is the coefficient of variation to represent the within-study variance that is not accounted for by other variables in our model. We have set $\tilde{\mu}$ to 0.1, i.e. assuming the within-study variation in random effects is likely to be very small relative to the between-study variation. The likelihood $L(x_{ij}|a_{ij}, b_{ij}, n_{ij})$ is calculated using the same formula as above, but in addition we calculate the likelihood of $q_i$ being the random effect adjustment for study $i$:

$$L(q_i|a, b_i) = \frac{(A_i(t_i)q_i)^{a_i}(1 - A_i(t_i)q_i)^{b_i}}{\mathrm{Beta}(a_i, b_i)}$$

##### 1.5.3 Likelihood function: drug volume data

Only three studies were identified that estimated antidepressant drug volumes: one for the private sector [83], one for the public sector in Gauteng province [115] and one for the public sector nationally [116]. In all three cases, the approach adopted to defining the likelihood is to estimate likely lower and upper bounds for the proportion of the population using antidepressants, and then to determine a normal likelihood based on these bounds. The approach to deriving the lower and upper bounds is further described below.

Table S7 summarizes the data from the private sector study. The published information (row A) is the number of person years of SSRI prescriptions in the private sector (values are approximate because they are read from a graph). We adjust this upward by dividing by 0.89, the proportion of private sector antidepressant users who are using SSRIs [90]. This adjusted total (B) is likely to be an upper bound on the true number of adults in medical schemes who are receiving antidepressants, for a number of reasons. Firstly, drug sales do not necessarily correspond to drugs used. Secondly, some of the drugs sold may be used for indications other than depression, and prescribed at dosages that would be considered sub-therapeutic for depression [76]. Thirdly, not all people receiving antidepressants through the private sector are medical scheme members. We obtain an upper bound on the private sector proportion by dividing the adjusted total (B) by the size of the adult medical scheme population (C, estimated from General Household Survey data). We obtain a lower bound by adjusting for the second and third sources of bias. We might conservatively assume that roughly half of people are receiving sub-therapeutic doses (it is conservative because the data are from the public sector [76], where there is more likely to be inappropriate prescribing/treatment for other indications [117]), and based on 2022 private sector data for antiretroviral treatment, we might estimate that roughly 82% of private sector drug use is by medical scheme members. The lower bound is thus calculated by multiplying the upper bound proportions by 0.41 ($0.5 \times 0.82$).

Table S7: Antidepressant prevalence in medical schemes

| | 2009 | 2010 | 2011 | 2012 | 2013 |
|---|---|---|---|---|---|
| Total individuals on SSRIs (A) | 290000 | 320000 | 325000 | 350000 | 390000 |
| Total individuals on antidepressants (B) | 325843 | 359551 | 365169 | 393258 | 438202 |
| Total adults in medical schemes (C) | 6505914 | 6831491 | 6362132 | 6971188 | 7322597 |
| % of adults in medical schemes on AD | | | | | |
| Lower bound (D) | 2.1% | 2.2% | 2.4% | 2.3% | 2.5% |
| Upper bound (E) | 5.0% | 5.3% | 5.7% | 5.6% | 6.0% |

AD = antidepressants, SSRIs = selective serotonin reuptake inhibitors

The public sector data from Gauteng are from a pharmaceutical database in 2017-18, focused on mental, neurological and substance use disorders [115]. Over the 2017-18 year, 16.25 million patient days of antidepressant treatment were prescribed, of which 9.97 million were for SSRIs. Converted to annual numbers, these correspond to 44 534 and 27 317 on antidepressants and SSRIs respectively. Much of the gap between these two figures is accounted for by amitriptyline, the most commonly used antidepressant. However, amitriptyline is frequently used for other conditions at sub-therapeutic doses [117], and may be used in conjunction with other antidepressants; SSRIs, on the other hand, are more likely to be used in treating depression, and it is unlikely that a single patient would be treated with multiple SSRIs. Thus the 44 354 and 27 317 might be considered upper and lower bounds respectively on the number of individuals receiving antidepressant treatment in the public sector. Two other sources of bias need to be considered. Firstly, Gauteng is a relatively wealthy province with better access to mental health care; in a national survey the average mental health expenditure per uninsured individual was 38% higher in Gauteng than nationally [118]. Secondly, people who are not medically insured might nevertheless access antidepressant treatment from the private sector, and if our denominator is the population that is not medically insured, it would be consistent to include these. One might roughly estimate an upper bound on this uninsured population accessing antidepressants through the private sector by taking the upper bound of 438 202 people on antidepressants in the private sector nationally (Table S7), multiplying by

0.18 (the proportion who are not insured) and then multiplying by 0.46 (the proportion of uninsured private sector ART use in 2022 that is in Gauteng), to get a total of 36 288. We thus obtain a lower bound on the prevalence of antidepressant use nationally, in the uninsured population, by dividing by 1.38, and then dividing by 7 968 465 (the adult population of Gauteng in 2017 who are not medical scheme beneficiaries, as estimated from the 2017 General Household Survey), which gives 0.25%. We obtain an upper bound by adding the public and private use of antidepressants in Gauteng (44 354 + 36 288), and dividing by the same denominator, which gives 1.01%.

We follow a similar approach in calculating of the lower and upper bounds for the other public sector data source (Table S8). This data set differs from the previous in that it contains only numbers receiving fluoxetine or citalopram (the most commonly used SSRIs), per 100 000 uninsured population, making it a lower bound on prevalence of antidepressant use. We derive an upper bound by adjusting for two likely sources of bias. Firstly we multiply by 1.63, the ratio of total antidepressant use to SSRI use in Gauteng (the same data source described in the previous paragraph). The adjustment of 1.63 is an upper bound because, as noted previously, many people receiving other antidepressants are receiving them at sub-therapeutic doses (for other indications) or are receiving them together with SSRIs (i.e. there could be double-counting of people on antidepressants). Secondly, we multiply by 1.81, the ratio of total antidepressant use by uninsured people in Gauteng to that secured through the public sector (calculated as 1 + 36288/44534, from the previous paragraph). This is also likely to be an upper bound, as Gauteng has a relatively large population who are uninsured but paying for healthcare privately.

Table S8: Antidepressant use in the public sector

| | 2019 | 2020 | 2021 | 2022 | 2023 |
|---|---|---|---|---|---|
| Lower bound | 0.34% | 0.47% | 0.39% | 0.38% | 0.46% |
| Upper bound | 1.00% | 1.40% | 1.14% | 1.12% | 1.36% |

Having obtained upper and lower limits for the $i^{\text{th}}$ 'data point', $u_i$ and $l_i$ respectively, we calculate the corresponding mean and standard deviation of the normal distribution, assuming that the lower and upper limits correspond to the 2.5 and 97.5 percentiles respectively of the normal distribution:

$$m_i = (l_i + u_i)/2$$
$$s_i = (u_i - l_i)/(2 \times 1.96)$$

We then calculate the model estimate of the prevalence of antidepressant use in health sector $h$ in year $t$ using a formula similar to that presented previously:

$$Z(h,t) = \frac{\sum_g \sum_c w_{g,t}(h,c) B(g,t) U^h \chi^c}{\sum_g \sum_c w_{g,t}(h,c)}$$

The normal likelihood for the $i^{\text{th}}$ data point is then

$$L(m_i | Z(h_i,t_i), s_i) = \frac{1}{\sqrt{2\pi} s_i} \exp\left(-\frac{\left(m_i - Z(h_i,t_i)\right)^2}{2{s_i}^2}\right)$$

##### 1.5.4 Posterior distributions

As with the calibration to depression prevalence data, we use Incremental Mixture Importance Sampling to numerically approximate the posterior distribution. The procedure is the same as that described in section 1.4.4, but the prior distributions are those specified in section 1.5.1, and the likelihood for each sampled parameter combination is calculated as the product of the likelihood functions defined in sections 1.5.2 and 1.5.3.

For the purpose of generating the final set of results, combining the uncertainty in the 10 depression incidence/recovery parameters and the uncertainty in the 10 antidepressant parameters, we randomly paired the 1000 parameter combinations in this step with the 1000 parameter combinations generated in the previous calibration (described in section 1.4.4). Since the parameters in each parameter set are mutually exclusive, the merged posterior parameter sample relates to 20 different parameters (10 + 10).

### 2. Additional results

#### 2.1 Comparison of prior and posterior distributions

Table S9 compares the prior distributions with the posterior distributions generated using the main model (Model A) and the models tested in the sensitivity analyses (Models B-E). Part a of the table focuses on the parameters varied in the calibration to depression prevalence data. Most of the posterior means in Model A are similar to the prior means, although there is a significantly smaller posterior estimate of the effect of excess COVID mortality on depression, and the posterior estimates of the accuracy of PHQ-9 and CES-D-10 are significantly lower than the prior means. Estimates of incidence rates are slightly higher than the prior means, while estimates of depression episode durations are slightly lower than the prior means (probably because it is difficult to match the odds ratios showing limited cross-survey associations with longer assumptions about depression duration). Posterior estimates tend to be similar across Models A-C and E, but in Model D the estimated average duration of depression episodes is significantly greater than in Model A. In addition, the annual incidence of depression in high-risk women, in Model A, is 0.21 (0.059/0.28), whereas the incidence rate in high-risk women is more than double in Model D (0.45).

Part b of the table focuses on the parameters varied in the calibration to antidepressant data. The posterior estimates for Model A are mostly similar to the prior means, although there is a high of the coefficient of variation in random effects, suggesting that almost all of the observed heterogeneity in antidepressant use across studies is not explained by the modelled covariates. There was also a relatively high posterior estimate of the effect of chronic diseases on antidepressant use, which suggests that it is difficult to reconcile the two antidepressant data sources (survey/cohort data versus drug distribution data) unless taking into account the over-representation of patients with chronic conditions among the former. Posterior estimates are similar across Models A-E.

Table S9a: Comparison of prior and posterior distributions (depression incidence and recovery)

| Parameter | Prior distribution (mean, 95% CI) | Posterior distribution (mean, 95% CI) Model A: Main model | Model B: Add time trend | Model C: No COVID effect | Model D: No low-risk depression | Model E: Larger high-risk |
|---|---|---|---|---|---|---|
| Mean duration of untreated symptoms | 9.00 (4.12-15.76) | 6.52 (5.63-7.43) | 6.37 (5.45-7.12) | 6.02 (4.97-7.00) | 14.7 (7.56-20.9) | 6.02 (5.25-6.91) |
| Incidence of depression in low-risk men | 0.027 (0.011-0.05) | 0.038 (0.034-0.044) | 0.039 (0.035-0.045) | 0.040 (0.035-0.047) | - | 0.035 (0.031-0.040) |
| Incidence of depression in low-risk women | 0.034 (0.013-0.064) | 0.059 (0.051-0.068) | 0.061 (0.054-0.069) | 0.062 (0.054-0.072) | - | 0.055 (0.048-0.062) |
| Ratio of incidence in low-risk group to high-risk group | 0.30 (0.02-0.75) | 0.28 (0.18-0.46) | 0.25 (0.19-0.33) | 0.23 (0.17-0.30) | - | 0.28 (0.22-0.37) |
| Increase in depression incidence per 10-year increase in age (RR) | 1.00 (0.81-1.21) | 1.05 (1.03-1.06) | 1.05 (1.04-1.06) | 1.05 (1.04-1.06) | 1.09 (1.06-1.11) | 1.05 (1.04-1.06) |
| % increase in depression incidence per unit excess deaths (per 100,000) | 0.039 (0.022-0.061) | 0.019 (0.015-0.025) | 0.016 (0.013-0.020) | - | 0.031 (0.02-0.041) | 0.018 (0.013-0.026) |
| Ratio of point prevalence to 12-month prevalence, using CIDI | 0.77 (0.60-0.91) | 0.89 (0.82-0.94) | 0.92 (0.88-0.95) | 0.89 (0.83-0.94) | 0.90 (0.82-0.96) | 0.89 (0.84-0.93) |
| Ratio of true prevalence to PHQ-9 prevalence | 0.35 (0.25-0.47) | 0.22 (0.20-0.24) | 0.21 (0.19-0.23) | 0.18 (0.16-0.20) | 0.23 (0.20-0.26) | 0.21 (0.19-0.24) |
| Ratio of true prevalence to CES-D prevalence | 0.35 (0.25-0.47) | 0.19 (0.17-0.21) | 0.19 (0.17-0.21) | 0.19 (0.17-0.22) | 0.20 (0.18-0.24) | 0.19 (0.17-0.21) |
| Effect of log(prevalence) on association between initial and later depression | -0.16 (-0.24,-0.08) | -0.18 (-0.23,-0.14) | -0.18 (-0.22,-0.14) | -0.17 (-0.25-,0.11) | - | -0.18 (-0.22,-0.13) |
| RR of depression incidence in 2025, relative to 2020 | 1.00 (0.27-2.19) | - | 0.95 (0.89-1.02) | - | - | - |
| Incidence of depression in high-risk men | 1.75 (0.04-6.46) | - | - | - | 0.21 (0.13-0.41) | - |
| Incidence of depression in high-risk women | 1.75 (0.04-6.46) | - | - | - | 0.45 (0.25-0.95) | - |

CES-D = Centre for Epidemiological Studies Depression Scale. CIDI = Composite International Diagnostic Interview, PHQ-9 = Patient Health Questionnaire-9, RR = relative rate.

Table S9b: Comparison of prior and posterior distributions (antidepressant use)

| Parameter | Prior distribution (mean, 95% CI) | Posterior distribution (mean, 95% CI) Model A: Main model | Model B: Add time trend | Model C: No COVID effect | Model D: No low-risk depression | Model E: Larger high-risk |
|---|---|---|---|---|---|---|
| Annual health seeking rate in women | 0.20 (0.05-0.44) | 0.27 (0.15-0.43) | 0.29 (0.16-0.50) | 0.29 (0.16-0.48) | 0.27 (0.14-0.46) | 0.27 (0.15-0.47) |
| RR of health seeking in men | 0.30 (0.08-0.66) | 0.41 (0.18-0.78) | 0.40 (0.16-0.75) | 0.39 (0.16-0.73) | 0.43 (0.12-0.85) | 0.41 (0.12-0.83) |
| Proportion prescribed antidepressants pre-2008 | 0.15 (0.02-0.36) | 0.25 (0.13-0.37) | 0.23 (0.11-0.40) | 0.24 (0.13-0.37) | 0.22 (0.10-0.37) | 0.25 (0.13-0.37) |
| RR of antidepressants in 2018 | 2.00 (1.00-3.34) | 2.79 (2.21-3.58) | 2.83 (2.13-3.63) | 2.85 (2.17-3.89) | 2.66 (2.00-3.58) | 2.72 (2.09-3.49) |
| RR of health seeking if previously treated | 9.00 (2.01-21.1) | 10.3 (3.04-21.6) | 10.0 (3.14-22.9) | 8.87 (2.95-18.2) | 10.5 (2.55-23.3) | 10.2 (3.77-21.4) |
| Average months on antidepressants while experiencing symptoms | 7.00 (2.41-14.0) | 7.43 (3.30-13.9) | 7.55 (3.43-14.9) | 7.40 (3.29-14.1) | 7.70 (3.79-13.3) | 7.28 (3.28-15.6) |
| Average years on antidepressants post-recovery | 6.00 (2.74-10.5) | 6.63 (3.70-10.7) | 6.73 (3.98-10.8) | 6.26 (3.82-9.97) | 6.75 (3.44-11.3) | 6.38 (3.44-10.5) |
| RR of antidepressant use in public sector | 0.09 (0.04-0.16) | 0.08 (0.08-0.09) | 0.09 (0.08-0.10) | 0.09 (0.08-0.10) | 0.08 (0.08-0.09) | 0.08 (0.08-0.09) |
| RR of antidepressants in people receiving treatment for other chronic conditions | 3.00 (1.37-5.25) | 4.92 (3.25-6.96) | 5.25 (3.43-7.49) | 4.95 (3.33-7.14) | 5.03 (3.25-7.23) | 5.08 (3.16-7.50) |
| Coefficient of variation in random effects | 0.50 (0.03-0.98) | 0.97 (0.91-1.00) | 0.97 (0.88-1.00) | 0.97 (0.89-1.00) | 0.96 (0.90-1.00) | 0.97 (0.90-1.00) |

RR = relative rate.

#### 2.2 Model calibration to age- and sex-specific depression prevalence data

Figure S7 shows the model calibration to the age- and sex-specific prevalence data. Although most of the model estimates are consistent with the survey data, it is notable that across most age and sex combinations, the SASH survey (conducted over 2002-2004) and 2010 National Income Dynamics survey measured a slightly lower depression prevalence than the model estimates.

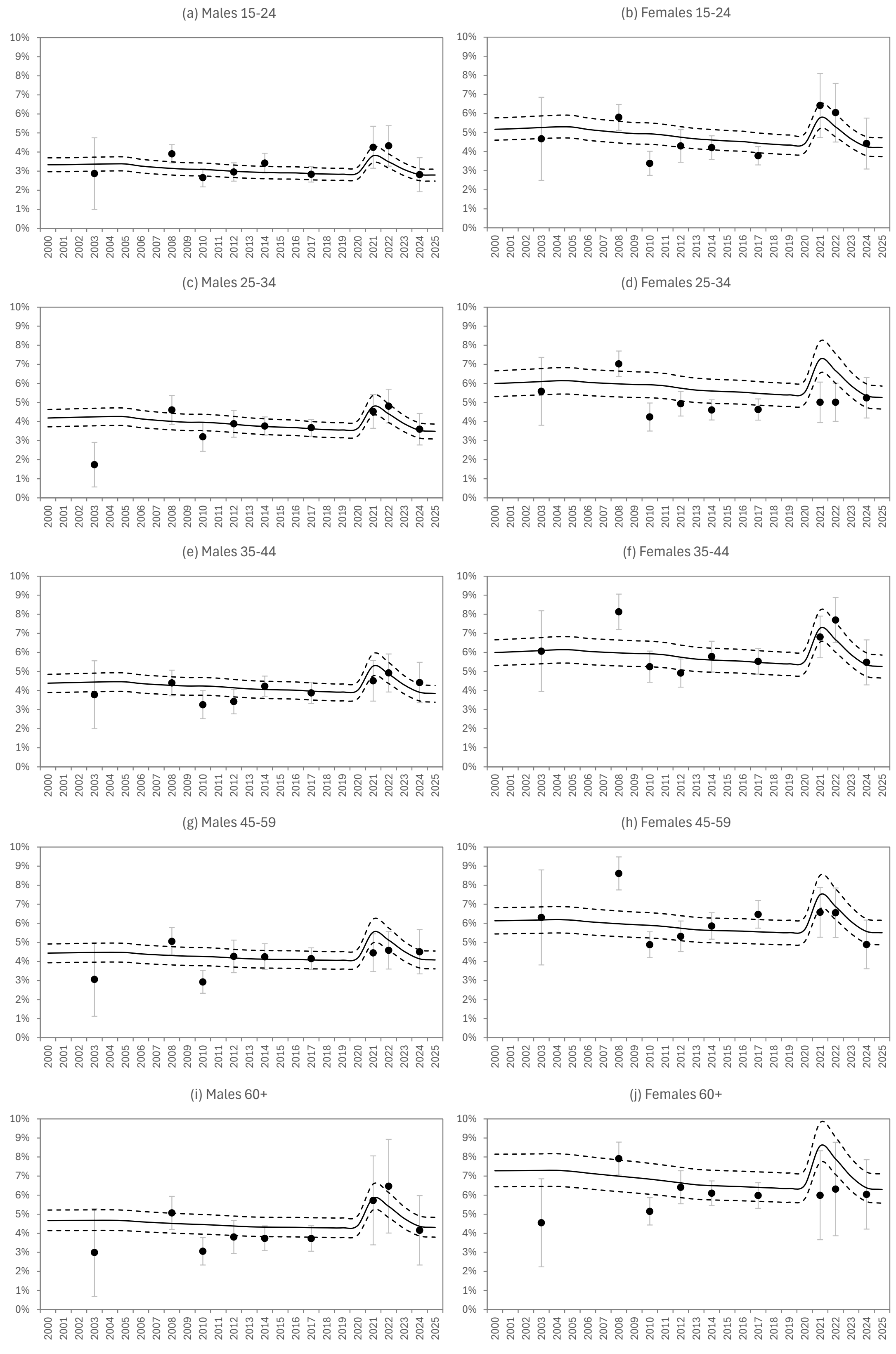

Figure S7: Trends in depression prevalence, by age and sex (Model A)

Solid lines represent posterior means of model estimates, dashed lines represent posterior 95% confidence intervals, and dots represent calibration data. Survey estimates have been adjusted downward by the posterior estimates of the correction factors (see Table S9).

#### 2.3 Inter-survey associations between depression (Model D)

Figure S8 shows the results from Model D (assuming no depression incidence in the high-risk group), comparing the results from the NIDS longitudinal surveys (in terms of the odds ratio relating depression in each survey to depression in the next survey) with the corresponding model results. The model results imply extremely high levels of association between initial and later depression, with odds ratios of around 20, i.e. the odds of depression in someone who was depressed in the previous survey (approximately two years previously) are roughly 20 times those in people who were not depressed in the previous survey. However, these model estimates are not consistent with the empirical estimates, which suggest odds ratios of around 1.5.

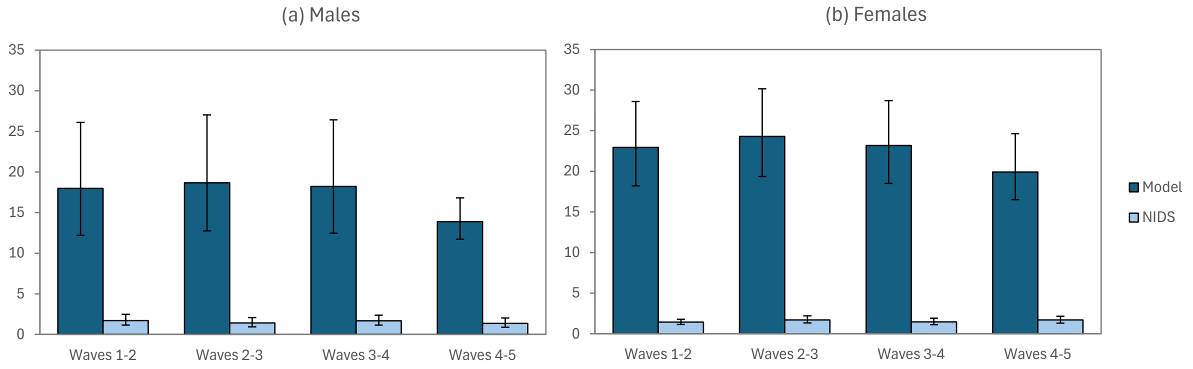

Figure S8: Odds ratios for association between initial and later depression

### Appendix A: Analysis of Quality of Life survey data

The Quality of Life survey is a household survey that has been conducted in the Gauteng province, at regular intervals since 2009. In the most recent survey, conducted over 2020-21, the survey questionnaire was extended to include questions about exposure to violence. The survey data have previously been analysed to assess the effect of different violence exposures on depression symptoms in sexual and gender minority groups, and showed that having experienced non-partner violence in the last year was associated with increased prevalence of depression symptoms (aOR 1.84, 95% CI: 1.24-2.73) [19]. However, this study did not assess the association between violence exposure and depression symptoms in the broader population, and did not assess the other correlates of violence exposure – questions which we attempt to address here.

**Methods**

The survey was based on a stratified random sample: within each district municipality (stratum), all wards were sampled (the primary sampling units), and within each ward, enumeration areas (EAs) were selected on a probability proportional to size basis. Within each EA, a random sample of households were sampled. All household members aged 18 and older were eligible to be interviewed. Questionnaires were administered by trained interviewers in face-to-face interviews. However, due to the sensitivity around questions about exposure to violence and sexual abuse/rape, respondents were asked to complete these questions themselves on handheld tablets. A more detailed description of the survey is provided elsewhere [119]. The survey data are freely available online, and can be accessed through the DataFirst facility (https://www.datafirst.uct.ac.za/).

Symptoms of depression were assessed using the PHQ-2 screening tool [120]. This is based on two questions about recently experiencing lack of interest/pleasure in doing things, and recently feeling down, depressed or hopeless. For the purpose of this analysis, we classify people as having depression symptoms if they had a PHQ-2 score of 2 or more (equivalent to reporting either symptom at least half the time, or both symptoms on "a few days").

Respondents were classified as having experienced violence if they reported having been hit, kicked, threatened or hurt with a weapon, in the last 12 months.

Multivariable logistic regression models were used to assess key predictors of depression and violence exposure. All analyses were conducted in STATA 17.0 and included survey weights and survey design specification.

**Results: Age and sex differences in violence exposure**

16% of adults in Gauteng reported having experienced violence in the last year, with the proportion being higher in men than in women (20% versus 12%) and higher among young adults [119]. Table A1 shows the effect of age and sex on exposure to violence in our multivariable model. There is a strong negative association between age and exposure to violence, and the effect of age appears to be relatively linear, with no improvement in model fit when comparing a quadratic age effect model (Model 1) to a linear age effect model (Model 2).

Table A1: Effects of age and sex on experience of violence

| | Model 1: aOR (95% CI) | Model 2: aOR (95% CI) |
|---|---|---|
| Sex | | |
| Males | 1 | 1 |
| Females | 0.56 (0.49-0.63) | 0.55 (0.49-0.63) |
| Per year of age | 0.982 (0.953-1.011) | 0.961 (0.957-0.966) |
| Per year of age$^2$ | 0.9997 (0.9994-1.0001) | - |

**Results: Effect of violence exposure on prevalence of depression**

Overall, 14% of respondents had symptoms of depression, and depression symptoms were correlated with reports of intimate partner violence, rape, childhood sexual abuse and non-partner violence [119]. Table A2 shows the most significant predictors of depression symptoms in a full multivariable model. Consistent with previous South African studies, we find a significantly higher prevalence in women compared to men, and in older adults compared to younger adults. Depression symptoms are also significantly associated with lower socio-economic status and recent experience of violence. Controlling for other forms of abuse (childhood sexual abuse and forced sex in the last year) did not substantially alter the association between recent experience of violence and depression symptoms.

Table A2: Predictors of depression symptoms in adults

| | Model 1: aOR (95% CI) | Model 2: aOR (95% CI) |
|---|---|---|
| Recent violence | 1.56 (1.38-1.75) | 1.46 (1.30-1.65) |
| Female sex | 1.52 (1.38-1.67) | 1.49 (1.36-1.64) |
| Age (ref. 18-24): | | |
| 25-34 | 1.22 (1.04-1.42) | 1.21 (1.03-1.41) |
| 35-44 | 1.45 (1.24-1.71) | 1.45 (1.23-1.70) |
| 45-59 | 1.44 (1.22-1.69) | 1.43 (1.22-1.68) |
| 60+ | 1.37 (1.16-1.62) | 1.39 (1.17-1.64) |
| Education (ref. none/primary) | | |
| Incomplete secondary | 0.88 (0.75-1.04) | 0.88 (0.75-1.05) |
| Completed secondary | 0.83 (0.68-1.01) | 0.84 (0.69-1.02) |
| Tertiary | 0.79 (0.64-0.98) | 0.79 (0.64-0.99) |
| Private health care | 0.60 (0.53-0.69) | 0.61 (0.53-0.70) |
| Employed | 0.74 (0.67-0.83) | 0.74 (0.67-0.83) |
| Childhood sexual abuse | | 1.12 (0.79-1.59) |
| Forced sex in last year | | 1.41 (1.23-1.62) |

### Appendix B: Sensitivity and specificity of depression screening tools

It may appear natural to adjust model estimates of the prevalence of depression using standard sensitivity and specificity estimates for the purpose of comparisons against surveys in which common screening tools have been used. However, this can lead to implausibly small differences across age and sex categories in the adjusted prevalence estimates. For example, suppose the modelled prevalence of depression is 3% in men and 6% in women (i.e. the modelled 'true' prevalence in women is double that in men). If we assume that a sensitivity of 83% and a specificity of 78% for the CES-D screening tool [6], the adjusted prevalence estimates would be 23.8% for men ($0.03 \times 0.83 + (1 - 0.03) \times (1 - 0.78)$) and 25.7% for women – only a very modest difference, proportionally. Survey data suggest much greater heterogeneity in depression prevalence across sub-populations, and this in turn suggests that it is unrealistic to assume that sensitivity and specificity parameters are the same across all sub-populations.

Rather than attempting to adjust the sensitivity and specificity parameters to take account of likely heterogeneities, we propose a simpler multiplicative adjustment to the model estimate, and assess how well this adjustment is likely to perform. We start by examining data on the relationship between the PHQ-9 screening score and depression, and then propose a simplified model based on these data, which is then used to assess the validity of the proposed multiplicative adjustment under different scenarios. The same principles are assumed to apply to other screening tools, such as CES-D-10.

The data come from a study that was conducted at three health facilities in Cape Town, to determine the prevalence of mental disorders and their correlates, as well as the performance of various diagnostic tools [59]. Health facility attenders were screened using the PHQ-9 tool and were also administered the MINI diagnostic interview (which we treat as the 'true' determination of whether people had depression). The PHQ-9 screening tool produces a score, from 0-27, indicating the severity of depression symptoms (a cutoff score of 10 is usually used in identifying "probable depression"). Figure B1 shows, for each possible level of the PHQ-9 score, the proportion of patients who had depression (based on the MINI diagnostic tool). The data suggest a roughly sigmoidal relationship, with steep increases in the probability of depression between scores of 10 and 20, and a stabilization of depression prevalence around 65-70%, at scores above 20.

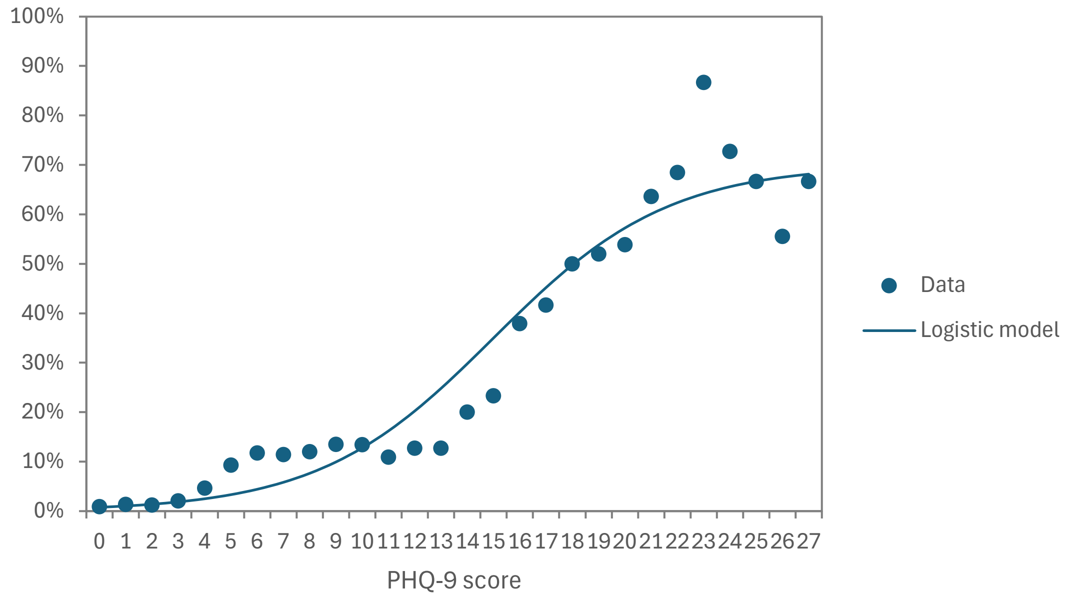

Figure B1: Prevalence of depression at different PHQ-9 scores
A moving average filter is applied to the data to reduce some of the stochastic noise in the data (for each value of $x$, we take the average over the range $x - 1$ to $x + 1$).

This relationship can be modelled using a logistic function of the form

$$p(x) = \frac{\alpha}{1 + \exp\big(-\theta(x - m)\big)}$$

where $p(x)$ is the probability that a person with PHQ-9 score of $x$ has depression, $\alpha$ is the maximum depression prevalence, $\theta$ is the logistic shape parameter and $m$ is the PHQ-9 score at the logistic function inflection point. Figure B1 shows that we obtain an adequate fit to the Cape Town study data when we set $\alpha$ to 70%, $\theta$ to 0.3 and $m$ to 15. We use this logistic function as the model in the evaluations that follow.

Suppose $f(x)$ represents the frequency of score $x$ in a particular population (or sub-population). Then the true prevalence of depression in the population is

$$\pi = \sum_{x=0}^{27} f(x)p(x)$$

The measured prevalence of 'probable depression', based on the standard PHQ-9 score of 10 or higher, is

$$\rho = \sum_{x=10}^{27} f(x)$$

Then the ratio of the true prevalence to the measured prevalence is $\pi/\rho$. The sensitivity of the PHQ-9 in detecting depression is

$$Se = \frac{1}{\pi} \sum_{x=10}^{27} f(x)p(x)$$

and the specificity is

$$Sp = \frac{1}{1-\pi} \sum_{x=0}^{9} f(x)(1-p(x))$$

In Table B1 we calculate these parameters for a number of hypothetical distributions of $f(x)$:

1. A gamma distribution with a mean of 7.22 and standard deviation of 4.35 (based loosely on the distribution of CES-D-10 scores in the 2008 NIDS survey [121], scaled to match the slightly maximum score for the PHQ-9).
2. A gamma distribution with a mean of 7.22 and standard deviation of 3.00 (lower variance)
3. A gamma distribution with a mean of 7.22 and standard deviation of 6.00 (higher variance)
4. A gamma distribution with a mean of 6.76 and standard deviation of 6.67 (estimated from the Cape Town study data in Figure B1 [59])
5. The actual distribution of depression scores in the Cape Town study (with no fitting of a gamma distribution)
6. A gamma distribution with a mean of 5.00 and standard deviation of 4.93 (lower mean than in the Cape Town study, but the same coefficient of variation)
7. A gamma distribution with a mean of 8.00 and standard deviation of 7.89 (higher mean than in the Cape Town study, but the same coefficient of variation)

(Although the gamma distribution is a continuous distribution and the $f(x)$ distribution is defined only for integer values of $x$, we approximate these by calculating cumulative probabilities at half-integer durations.)

Table B1: Performance of PHQ-9 screening tool (relative to the true prevalence of depression) under different assumptions about $f(x)$

| Distribution, $f(x)$ | 1 | 2 | 3 | 4 | 5 | 6 | 7 |
|---|---|---|---|---|---|---|---|
| Mean of $f(x)$ | 7.22 | 7.22 | 7.22 | 6.76 | 6.76 | 5.00 | 8.00 |
| Standard deviation of $f(x)$ | 4.35 | 3.00 | 6.00 | 6.67 | 6.67 | 4.93 | 7.89 |
| True depression prevalence, $\pi$ | 9.8% | 8.1% | 11.4% | 11.0% | 12.5% | 6.8% | 13.9% |
| Measured prevalence, $\rho$ | 25.0% | 20.4% | 26.8% | 24.5% | 29.9% | 14.8% | 30.6% |
| Ratio: true to measured, $\pi/\rho$ | 0.391 | 0.399 | 0.425 | 0.447 | 0.418 | 0.460 | 0.455 |
| Sensitivity, $Se$ | 67.4% | 50.6% | 77.5% | 79.2% | 84.7% | 66.0% | 84.2% |
| Specificity, $Sp$ | 79.6% | 82.3% | 79.7% | 82.2% | 78.0% | 88.9% | 78.1% |
| False positive rate, 1 – $Sp$ | 20.4% | 17.7% | 20.3% | 17.8% | 22.0% | 11.1% | 21.9% |

Table B1 shows that across these seven different distributions, the ratio $\pi/\rho$ is relatively stable, varying between 0.39 and 0.46 (average 0.43). In contrast, the sensitivity is highly variable, ranging from 51% to 85%, and the false positive rate (1 – specificity) varies between 11% and 22%). This confirms that it is more reasonable to assume stability of the $\pi/\rho$ ratio than it is to assume stability of the sensitivity and specificity parameters.

Although the ratio $\pi/\rho$ is stable at around 0.43 in this example, it should not be assumed that 0.43 is necessarily close to the 'true' ratio for PHQ-9. For example, if we were to replace the value of $\alpha$ with 0.35 (half of the previous value), the ratio would instead be stable at around 0.21. If we were to set the PHQ-9 threshold for 'probable depression' at 8 instead of 10 (as recommended in one South African study [101]), the average ratio would instead be around 0.30 (with less stability).
